# Multidimensional socioeconomic gradients in biological age acceleration across 12 countries in the SHARE study

**DOI:** 10.64898/2026.09.11.26361838

**Authors:** Haotian Guo

## Abstract

**Background:** Socioeconomic patterning has been documented across epigenetic, allostatic-load and clinical-biomarker measures of biological ageing. How these gradients vary across dimensions of social position and national settings remains unclear.

**Methods:** Cross-sectional analyses used 2015 data from the Survey of Health, Ageing and Retirement in Europe (SHARE) on 10,043 adults aged 50–75 years in 12 countries, with health follow-up through Wave 9. Sex-calibrated Klemera–Doubal method (KDM) age acceleration combined nine physiological indicators. Survey-weighted models examined four stand-alone indicators of socioeconomic position—respondent education, highest parental education, wealth and income—alongside education-based relational specifications for educational mobility and jointly modelled parental and respondent education ranks; *τ* summarised between-country dispersion.

**Results:** Across SES representations, six of seven pooled coefficients were inverse, ranging from *−*0.067 SD for highest parental education to *−*0.221 for wealth rank, while mutually adjusted parental education rank was close to zero (*β* = *−*0.017). A common-sample omnibus test showed overall variation among the seven specification-specific coefficients (*P <* 0.001). Country-specific point estimates were predominantly negative. Estimated between-country dispersion was larger for respondent education, income, educational mobility and mutually adjusted respondent education rank (*τ* = 0.053– 0.074), but small or at the boundary for highest parental education, wealth and mutually adjusted parental education rank (*τ* = 0.000–0.013).

**Conclusions:** Socioeconomic gradients in KDM age acceleration were predominantly inverse but varied in magnitude across representations of socioeconomic position; their direction was broadly shared across countries, while the extent of between-country heterogeneity varied across representations.

**Key Messages:**

- Socioeconomic gradients in KDM age acceleration were predominantly inverse, but their magnitude varied across representations of socioeconomic position.
- These gradients showed substantial cross-national commonality in direction, alongside measure-specific differences in between-country heterogeneity.
- KDM age acceleration predicted subsequent health outcomes, while the pooled socioeconomic pattern was reproduced across alternative weighting and biomarker specifications.

## 1 Background

Socioeconomic inequalities in later-life health may become biologically embedded through cumulative differences in material resources, psychosocial stress, occupational and environmental exposures, health behaviour and access to care [1]. Consistent with this perspective, socioeconomic gradients have been documented across multiple measures of biological ageing, including epigenetic ageing, allostatic load, clinical-biomarker age and proteomic profiles [2–6]. Social, inequality-related, and financial stress have also been found to be associated with epigenetic ageing measured by DNA-methylation clocks [7]. Among clinical-biomarker approaches, Klemera–Doubal method (KDM) biological age summarises multisystem physiological profiles relative to age-related reference patterns [8, 9]. Educational inequalities have been documented in KDM age, with evidence of widening gradients across successive US samples [10]. However, whether socioeconomic patterning of biological ageing differs across dimensions of social position and whether these gradients are broadly shared across national settings remain less clear.

One reason to expect variation is that socioeconomic position is multidimensional and unfolds across the life course. Parental education reflects early-life social origins, respondent education later attained position, income current resource flows, and wealth accumulated resources and the capacity to buffer shocks. Intergenerational educational patterns add information about adult attainment relative to parental background rather than either generation in isolation. Recent longitudinal evidence further suggests that socioeconomic conditions at different life-course stages may be relevant to biological ageing: both childhood and adulthood socioeconomic conditions have been associated with DNA methylation-based measures, with adulthood conditions partially mediating associations with childhood conditions [11]. In SHARE, parental education has been associated with later-life C-reactive protein [12], while research on educational mobility shows that estimated associations depend on how origin, destination and mobility are conceptualised and parameterised [13–16]. Together, these distinctions make it substantively important to ask whether socioeconomic gradients in biological ageing differ in magnitude across alternative representations of socioeconomic position.

A second question is whether these gradients are broadly shared across countries or shaped by national context. Welfare provision, pension protection, healthcare financing and labour-market conditions may affect both exposure to disadvantage and the resources available to buffer its consequences [17–19]. European studies document cross-national variation in mortality and healthy-ageing inequalities [20–22], but direct comparative evidence for KDM age acceleration remains limited. SHARE therefore provides an opportunity to distinguish cross-national commonality in the direction of socioeconomic gradients from variation in their magnitude, and to examine whether between-country differences are patterned by broader institutional and socioeconomic conditions.

A further issue concerns the health relevance of the physiological variation captured by KDM age acceleration. KDM measures have prospectively predicted mortality, disability and other age-related outcomes [23–26]. Because biological age is a latent construct and biomarker composites differ in content [27, 28], associations with subsequent health outcomes provide complementary evidence that the variation indexed by a given KDM measure is consequential for later-life health.

Against this background, I examined whether socioeconomic gradients in KDM age acceleration varied across representations of socioeconomic position spanning social origin, educational attainment, material resources and intergenerational educational relations. I then assessed whether these gradients were broadly shared across 12 countries in SHARE or varied in magnitude between countries, whether they differed by sex and baseline age, and whether cross-national variation was patterned by welfare, pension, healthcare and broader socioeconomic conditions. Finally, I examined whether KDM age acceleration predicted subsequent health and whether the main socioeconomic patterns were robust to alternative weighting and biomarker specifications.

## 2 Methods

### 2.1 Data, participants and survey design

The Survey of Health, Ageing and Retirement in Europe (SHARE) follows adults aged 50 years or older and their partners across Europe and Israel [29]. Wave 6 (2015) supplied interviews, dried-blood-spot (DBS) biomarkers, grip strength and peak flow; Waves 7–9 supplied prospective health outcomes. Analyses used released interview, generated-variable, DBS and Gateway Harmonized SHARE data; DBS collection and processing are documented elsewhere [30, 31].

Of 24,218 DBS respondents, 18,063 were aged 50–75 years with valid sex and country and positive finite DBS weights. The upper age limit follows earlier KDM research [32]. The primary biomarker panel was complete for 10,043 respondents in Belgium, Denmark, Estonia, France, Germany, Greece, Israel, Italy, Slovenia, Spain, Sweden and Switzerland. SES models used specification-specific complete cases; parental-education models excluded Greece because only two primary-panel respondents had usable parental education. The reduced panel included up to 14,015 respondents.

Models used official DBS weights and released strata and primary sampling units (PSUs), nested within country and subsample as appropriate. Respondent-level PSUs substituted where released PSU information was insufficient, and survey estimation adjusted for single-PSU strata [33]. Kish effective sample size (ESS) summarised weight dispersion. Table 1 describes the primary sample before SES-specific exclusions; supplementary methods detail survey-design and support rules.

**Table 1.** Baseline characteristics of the primary-panel KDM age acceleration sample. Primary nine-indicator panel; Wave 6; ages 50–75. N = 10,043; 12 countries; Kish ESS = 1,186.

| Characteristic | Weighted mean (SD) |  |
| --- | --- | --- |
|  | Unweighted n (weighted %) | Unavailable n |
| **Demographic characteristics** |  |  |
| Age (years) | 61.65 (6.72) | 0 |
| Female | 5,798 (53.2%) | 0 |
| **Socioeconomic characteristics** |  |  |
| Respondent education (years) | 11.92 (4.50) | 0 |
| Highest parental education (years) | 8.80 (5.03) | 720 |
| Wealth rank (0–100) | 51.96 (28.89) | 40 |
| Income rank (0–100) | 51.71 (29.11) | 1,166 |
| Educational mobility (rank points) | 0.86 (26.32) | 720 |
| Parental education rank (0–100) | 51.38 (26.14) | 720 |
| Respondent education rank (0–100) | 51.52 (28.43) | 720 |
| **Country** |  |  |
| Belgium | 1,408 (4.3%) | – |
| Denmark | 1,589 (2.7%) | – |
| Estonia | 1,688 (0.6%) | – |
| France | 123 (20.2%) | – |
| Germany | 1,220 (31.5%) | – |
| Greece | 231 (2.9%) | – |
| Israel | 119 (0.7%) | – |
| Italy | 567 (18.1%) | – |
| Slovenia | 795 (0.8%) | – |
| Spain | 382 (11.4%) | – |
| Sweden | 1,047 (3.6%) | – |
| Switzerland | 874 (3.3%) | – |

DBS = dried blood spot; SD = standard deviation. Means, SDs and percentages use DBS weights and available observations. Counts (n) describe respondents. Country was required for sample inclusion; dashes indicate that missingness does not apply to country-category rows. Kish ESS denotes the Kish effective sample size, a summary of weight dispersion rather than parameter-specific precision. Country percentages describe shares of the weighted analytic sample.
Wealth and income refer to net wealth and annual after-tax income of the respondent and spouse/partner, if any. Their ranks are summarised separately within each of the five imputed datasets; the reported means and SDs are their respective averages. Socioeconomic status (SES) characteristics use their pre-standardisation scales; educational mobility is measured in residualised rank points. KDM = Kleméra–Doubal method. KDM age acceleration is standardised within sex and the primary panel. Ranked SES variables retain the eligible DBS reference distribution described in the supplementary methods.

### 2.2 KDM age acceleration

The primary nine-indicator panel comprised glycated haemoglobin, total haemoglobin, high-density lipoprotein cholesterol, total cholesterol, log triglycerides, log C-reactive protein, log cystatin C, peak flow and grip strength. A reduced five-indicator panel retained glycated haemoglobin, total cholesterol, log C-reactive protein, log cystatin C and grip strength, increasing complete-indicator eligibility. KDM parameters were estimated separately by sex and panel using DBS weights and a finite-age-range correction [8, 34]. Within sex, biological age was regressed on a three-degree-of-freedom natural age spline; residuals were standardised by their weighted SD. Higher scores indicate older biological age than expected for chronological age. Age acceleration therefore represents an age-adjusted cross-sectional physiological profile rather than within-person ageing rate. Each panel retained its own calibration and outcome scale.

### 2.3 SES measures and reference scales

Four stand-alone indicators were examined: respondent education, highest parental education, wealth rank and income rank. Two education-based relational specifications were examined alongside them. Educational mobility was the weighted regression residual of respondent education rank on parental education rank; a separate origin–destination model entered parental and respondent education ranks jointly, yielding two mutually adjusted coefficients. Thus, six specifications yielded seven coefficients.

Parental ISCED-97 categories were mapped to years, taking the highest available parental value; educational ranks were constructed within country and birth-cohort bands. Wealth represented net assets of the respondent and spouse or partner, if any. Income was their annual total after taxes and social contributions, including earnings, pensions, benefits, transfers, rent and investment income. Neither monetary measure was equivalised for household size. Both were reconstructed and ranked in each of SHARE’s five imputations; supplementary methods detail components and eligibility.

Fixed country-specific reference scales kept SES units constant across analytic samples and weighting schemes. Variables were centred and scaled using available values from 39,095 completed Wave 6 interviews at ages 50–75 with positive interview weights; monetary moments were calculated by imputation. Rank mappings were defined before panel-specific exclusions and then standardised using interview-based moments. Monetary models used identical participants across imputations and Rubin pooling with finite-sample degrees of freedom [35, 36].

### 2.4 Pooled associations and between-country variation

Survey-weighted Gaussian common-slope models fitted the four stand-alone indicators and educational mobility separately, adjusting for country, sex and a three-degree-of-freedom natural age spline; parental and respondent ranks were mutually adjusted in the joint model. The adjustment set comprised country, sex and age to characterise total socioeconomic patterning. Pooled coefficients are respondent-level common-slope summaries; SES-by-country interactions provided country-specific coefficients, and Wald tests assessed equality of country slopes.

Between-country dispersion was summarised by *τ*, the SD of country coefficients, estimated by restricted maximum likelihood (REML) using their full sampling covariance matrix. Joint ranks used separate variance components under a diagonal random-effects structure. Profile-REML intervals quantified uncertainty in dispersion; pooled coefficients remained those from the survey-weighted common-slope models.

Within each panel, a cross-SES common sample aligned all six specifications for an omnibus test of equality among the seven pooled coefficients, imposing six restrictions. Stacked survey covariance and finite-sample D1 pooling across five monetary imputations accounted for dependence between estimates [37, 38]. For the primary panel, pairwise contrasts were organised within the two conceptual sets: six differences among the four stand-alone indicators and three among the relational coefficients (Table 3). These used the same common sample, preserved cross-model survey covariance and used scalar Rubin pooling; Holm adjustment was applied jointly to the nine pairwise *P* values. Confidence intervals are pointwise 95% intervals; other tests use nominal two-sided *P* values. Inference conditions on estimated KDM calibration and SES reference-scale parameters.

### 2.5 Prospective health associations

Prospective outcomes were activities of daily living (ADL; six items) and instrumental ADL (IADL; nine items) limitations at Waves 7–9, Wave 9 cognition and mortality. Incident limitation was defined among respondents without the corresponding limitation at baseline. Cognition combined memory, verbal fluency, numeracy and orientation using fixed country-specific Wave 6 calibration. Logistic, Gaussian and Cox survey models adjusted for age, sex and country; functional and cognitive models also adjusted for follow-up interval, and cognition for baseline performance. Mortality follow-up ended at death or the last regular interview and was capped at seven years. Supplementary methods detail eligibility and date handling.

### 2.6 Additional heterogeneity and sensitivity analyses

Exploratory meta-regressions examined public social expenditure, disposable-income inequality, low-earner pension replacement, out-of-pocket health expenditure, GDP per capita and unemployment separately. Sex and baseline-age interactions were examined separately while retaining country-varying SES slopes. Age interactions used linear and natural-spline terms; age-specific coefficients were common to both sexes. Weighting analyses compared official DBS weights, within-country trimmed DBS weights, unit weights, stabilised inverse-probability-of-selection (SIPW) weights and trimmed SIPW weights. Selection models targeted complete primary-panel measurements among eligible DBS respondents using demographic, SES and DBS-collection variables and were re-estimated in 1,000 survey bootstrap replicates. Reduced-panel analyses used panel-specific and within-SES common-panel samples without recalibrating either outcome. Supplementary methods provide full specifications and inferential details.

### 2.7 Software

Analyses were conducted in R using `survey, splines, survival, metafor, clubSandwich and wildmeta` [33, 39–41]. Bootstrap *P* values used the plus-one convention [42].

## 3 Results

### 3.1 Sample characteristics

The primary sample included 10,043 respondents in 12 countries, with weighted mean age 61.65 years (SD 6.72), 53.2% women and mean education 11.92 years (Table 1). Kish ESS was 1,186. SES-specific samples ranged from 8,877 for income to 10,043 for respondent education; parental-education models included 9,321 respondents in 11 countries.

### 3.2 Pooled socioeconomic associations

All four stand-alone SES indicators were inversely associated with KDM age acceleration (Table 2): coefficients per reference SES SD were *−*0.099 SD (95% CI *−*0.160 to *−*0.038) for respondent education, *−*0.067 (*−*0.117 to *−*0.017) for parental education, *−*0.221 (*−*0.277 to *−*0.165) for wealth and *−*0.215 (*−*0.271 to *−*0.159) for income. In the common sample, wealth and income were each more inverse than respondent and parental education in all four Holm-adjusted contrasts (Holm *P* values *<* 0.001 to 0.0112). By contrast, differences within the education pair and within the material-resource pair were small: the respondent–parental education contrast was *−*0.053 SD (95% CI *−*0.110 to 0.003; *P*_Holm_ = 0.194), and the wealth–income contrast was *−*0.026 SD (*−*0.087 to 0.036; *P*_Holm_ = 0.407); Table 3).

**Table 2.** Pooled socioeconomic associations with primary-panel KDM age acceleration.

| SES indicator / coefficient | $N$ / countries | $\beta$ (95% CI) | $P(\beta = 0)$ |
| --- | --- | --- | --- |
| **Stand-alone SES indicators** |  |  |  |
| Respondent education | 10,043 / 12 | -0.099 (-0.160, -0.038) | 0.002 |
| Highest parental education | 9,321 / 11 | -0.067 (-0.117, -0.017) | 0.008 |
| Wealth rank | 10,003 / 12 | -0.221 (-0.277, -0.165) | <0.001 |
| Income rank | 8,877 / 12 | -0.215 (-0.271, -0.159) | <0.001 |
| **Education-based relational specifications** |  |  |  |
| Educational mobility | 9,321 / 11 | -0.121 (-0.165, -0.078) | <0.001 |
| Parental education rank (joint) | 9,321 / 11 | -0.017 (-0.064, 0.030) | 0.477 |
| Respondent education rank (joint) | 9,321 / 11 | -0.127 (-0.175, -0.079) | <0.001 |

**Estimates and samples:** Pooled coefficients from survey-weighted common-slope models are SD of KDM age acceleration per fixed within-country reference SD of SES. The two coefficients from the joint education-rank specification were mutually adjusted. Models adjust for baseline age (natural spline, 3 df), sex and country. Wealth and income combine five monetary imputations. CIs are pointwise, and component $P$ values are two-sided tests of each coefficient against zero. Displayed coefficients use specification-specific samples; the coefficient-equality test below uses a separate cross-SES common sample. Pooled coefficients are respondent-level common-slope summaries; country-specific slopes are reported separately. SES reference moments use completed Wave 6 interviews at ages 50–75 with positive interview weights (39,095 respondents overall), within country and monetary imputation. These moments remain fixed across outcomes, samples and weighting schemes; rank construction is unchanged. Supplementary methods give construction and eligibility details.

**Table 3.** Pairwise contrasts of pooled socioeconomic coefficients for primary-panel KDM age acceleration.

| Contrast | Coefficient difference | Pointwise 95% CI | Raw $P$ | Holm-adjusted $P$ |
| --- | --- | --- | --- | --- |
| **Stand-alone SES indicators** |  |  |  |  |
| Respondent education – parental education | −0.0533 | [−0.1097, 0.0032] | 0.0646 | 0.194 |
| Respondent education – wealth | 0.1213 | [0.0541, 0.1886] | < 0.001 | 0.00294 |
| Respondent education – income | 0.0956 | [0.0354, 0.1557] | 0.00186 | 0.0112 |
| Parental education – wealth | 0.1746 | [0.1094, 0.2398] | < 0.001 | < 0.001 |
| Parental education – income | 0.1488 | [0.0863, 0.2114] | < 0.001 | < 0.001 |
| Wealth – income | −0.0258 | [−0.0872, 0.0357] | 0.407 | 0.407 |
| **Coefficients from education-based relational specifications** |  |  |  |  |
| Educational mobility – parental education rank (joint) | −0.0989 | [−0.1729, −0.0249] | 0.00877 | 0.0351 |
| Educational mobility – respondent education rank (joint) | 0.0073 | [−0.0006, 0.0151] | 0.0689 | 0.194 |
| Parental education rank (joint) – respondent education rank (joint) | 0.1062 | [0.0297, 0.1827] | 0.0065 | 0.0325 |

**Estimates and inference:** All contrasts use the primary-panel cross-SES common sample of 8,252 participants from 11 countries. Differences are the first coefficient minus the second, in KDM age-acceleration SD per one fixed within-country reference SD of SES. Parental education denotes highest parental education; wealth and income denote total wealth percentile and relative income percentile, respectively. Educational mobility denotes residualised rank mobility. The two joint education-rank coefficients are mutually adjusted. Tests preserve survey-design cross-model covariance and use scalar Rubin pooling across five imputations with finite-sample degrees of freedom. All $P$ values are two-sided. Holm adjustment is applied jointly to all nine contrasts; confidence intervals are pointwise 95% intervals.

Among the relational coefficients, educational mobility (*β* = *−*0.121, 95% CI *−*0.165 to *−*0.078) and mutually adjusted respondent rank (*β* = *−*0.127, *−*0.175 to *−*0.079) were inverse, whereas mutually adjusted parental rank was close to zero (*β* = *−*0.017, *−*0.064 to 0.030). Mobility and respondent rank were each more inverse than parental rank (*P*_Holm_ = 0.0351 and 0.0325), while differing little from one another (difference = 0.007 SD, 95% CI *−*0.001 to 0.015; *P*_Holm_ = 0.194). Across all seven coefficients, the common-sample omnibus test provided evidence of overall variation (*P <* 0.001).

### 3.3 Between-country variation

Country-specific point estimates were predominantly negative, indicating broad cross-national commonality in direction: all 12 estimates were negative for wealth and 11 of 12 for respondent education and income (Table S6). The extent of between-country variation, however, differed across SES dimensions. Heterogeneity was limited for wealth (*τ* = 0.000; country-slope equality *P* = 0.564), parental education (*τ* = 0.013; *P* = 0.258) and mutually adjusted parental education rank (*τ* = 0.000; *P* = 0.354). By contrast, respondent education (*τ* = 0.074; *P <* 0.001), income (*τ* = 0.071; *P* = 0.001), educational mobility (*τ* = 0.053; *P* = 0.004) and mutually adjusted respondent education rank (*τ* = 0.057; *P* = 0.003) showed greater between-country variation (Figure 1).

**Figure 1.**
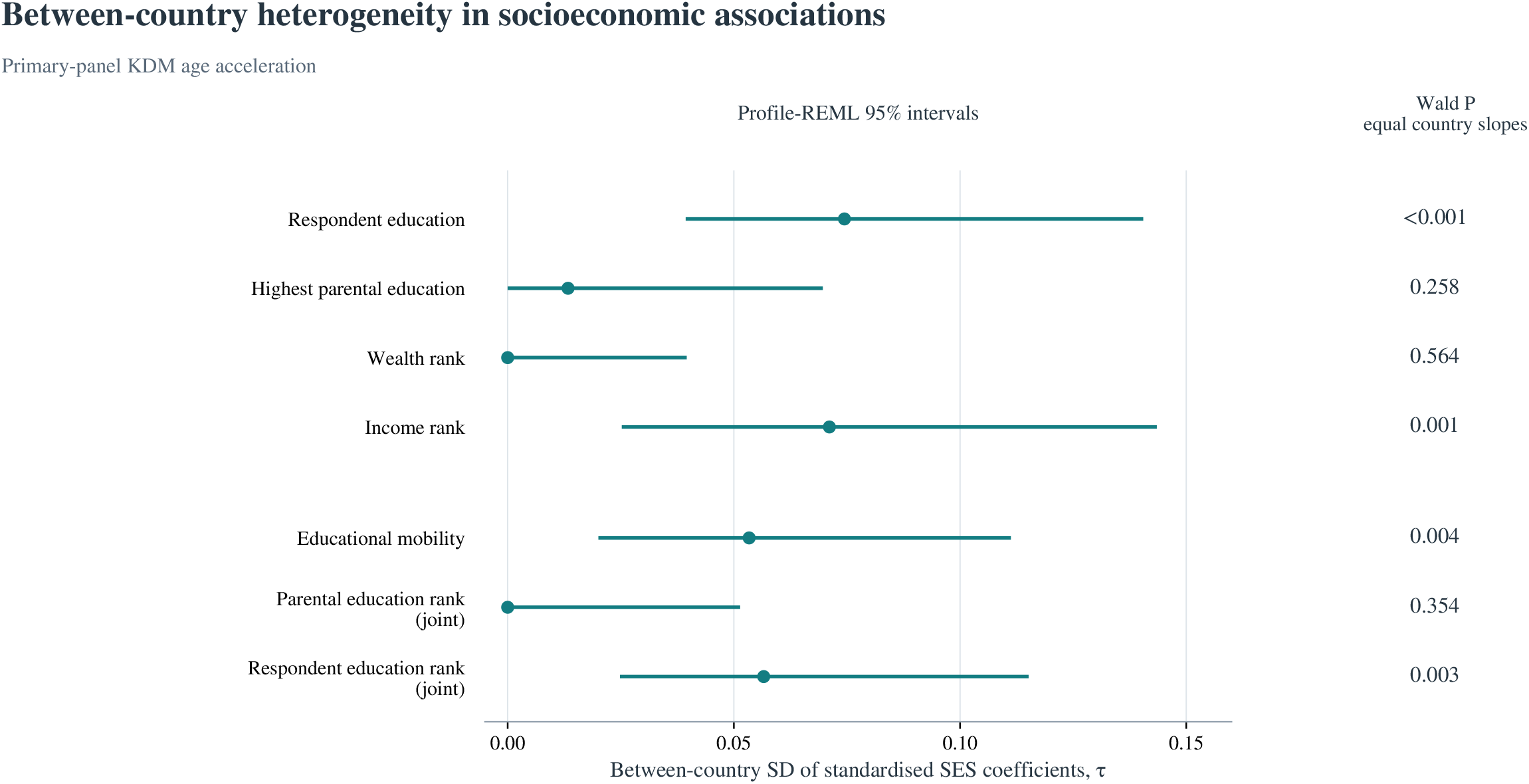
Between-country heterogeneity in socioeconomic associations. Between-country SDs (*τ*) of standardised country-specific SES coefficients for primary-panel KDM age acceleration. Each coefficient is measured in KDM age-acceleration SD per fixed within-country reference SES SD; *τ* uses the same coefficient units. On the left, horizontal lines show pointwise profile-REML 95% CIs for *τ*. On the right, Wald *P* values test equality of country slopes within each coefficient component. Educational mobility is residualised rank mobility; joint educational ranks are mutually adjusted. Pooled estimates and specification-specific samples appear in Table 2; country coefficients appear in Table S6. The joint country-slope test for both educational ranks was *F* (20, 6165) = 1.991, *P* = 0.005 (*N* = 9,321; 11 countries). **Alt text:** A horizontal plot displays between-country coefficient SDs and pointwise profile-REML intervals for seven specification-specific SES coefficients, visually grouped as four stand-alone indicators and three coefficients from education-based relational specifications. Respondent education and income have the largest point estimates. Wealth and mutually adjusted parental rank have boundary estimates. A separate right-hand column gives Wald P values for equal country slopes.

### 3.4 Prospective health associations

Higher acceleration predicted subsequent functional limitations, poorer cognition and mortality (Table S1). Wave 9 odds ratios per SD were 1.513 (95% CI 1.196–1.913) for incident ADL and 1.467 (1.143–1.881) for incident IADL limitation, with the same direction at Waves 7 and 8. The cognition coefficient was *−*0.075 reference SD (*−*0.140 to *−*0.010), conditional on baseline cognition. The mortality hazard ratio was 1.919 (1.647–2.236), based on 9,219 participants and 560 deaths; median follow-up was 6.67 years.

### 3.5 Additional heterogeneity and sensitivity analyses

Country-context estimates were generally imprecise across the small number of countries, and showed no clear pattern linking between-country variation in socioeconomic gradients to the examined welfare, pension, healthcare or macroeconomic characteristics (Tables S4 and S7). Sex- and baseline-age-specific estimates showed no consistent pattern of heterogeneity (Table S3). Pooled associations were consistently inverse across all five weighting schemes (Figure 2). SIPW estimates were close to those using official DBS weights, whereas unit weighting produced somewhat larger changes in magnitude and precision; weighting diagnostics and bootstrap support are reported in Table S8. Reduced-panel analyses also reproduced the main pooled pattern in both within-SES common-panel and panel-specific samples (Table S2). All seven pooled point estimates remained negative; wealth and income were again among the largest inverse coefficients, while mutually adjusted parental education rank remained close to zero. The cross-SES common-sample omnibus test again showed variation among the seven coefficients (*P <* 0.001), and prospective associations retained the same directions.

**Figure 2.**
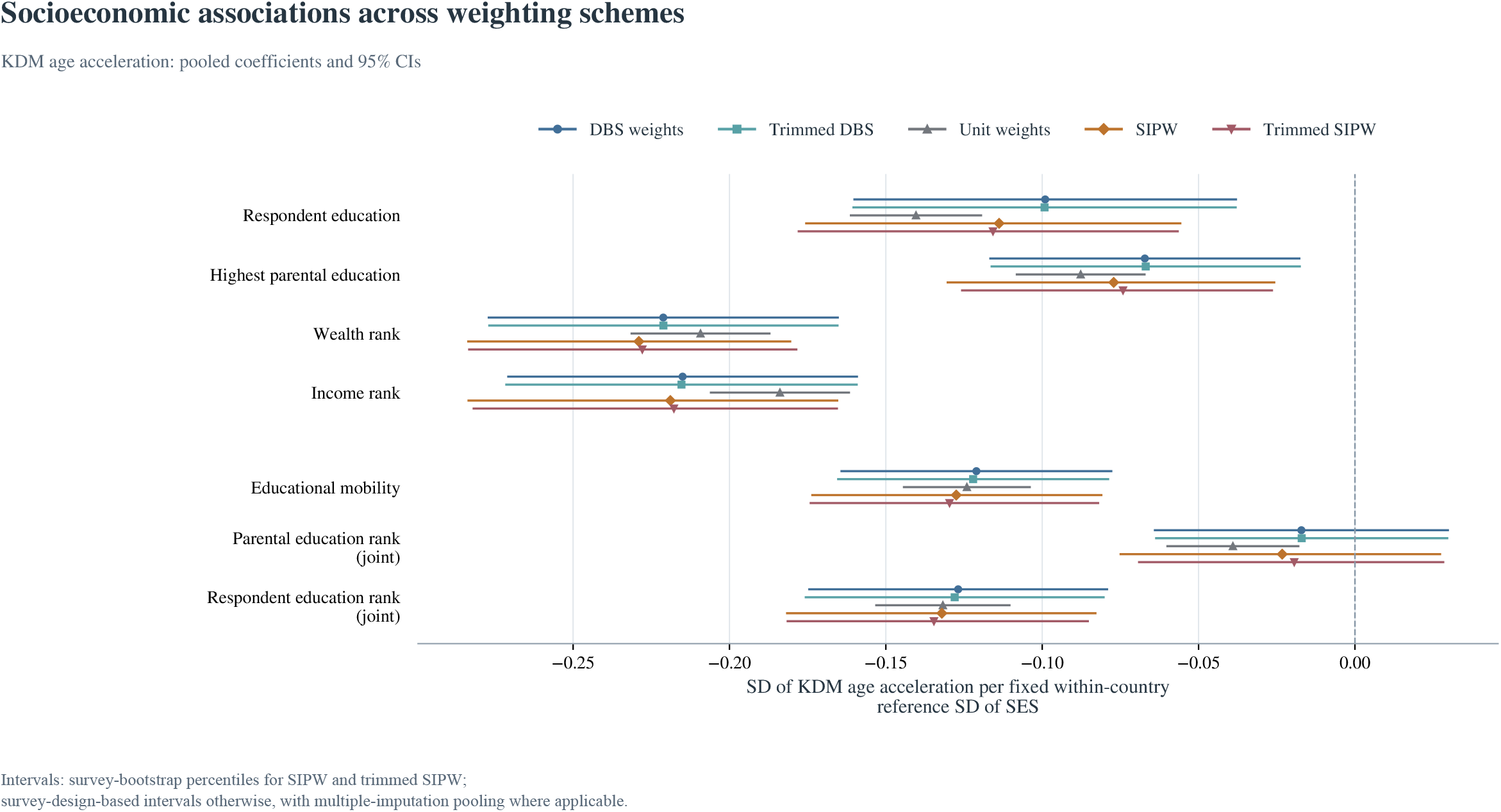
Socioeconomic associations across weighting schemes. Pooled primary-panel coefficients and pointwise 95% CIs under five weighting schemes, expressed in outcome SD per fixed within-country reference SD of SES. DBS denotes dried-blood-spot weights; SIPW denotes stabilised inverse-probability-of-selection weighting for complete primary-panel measurements among eligible DBS respondents. SIPW and trimmed SIPW intervals are survey-bootstrap percentile intervals from 1,000 draws with selection weights re-estimated; other schemes use survey-design-based intervals, with multiple-imputation pooling where applicable. Within-country 1st/99th-percentile caps apply to DBS weights or the SIPW selection multiplier, respectively. Educational mobility is residualised rank mobility; joint educational ranks are mutually adjusted. Participants and outcome and SES scales are held constant across weighting schemes within each specification. Table S8 reports ESS and bootstrap completeness. **Alt text:** Seven coefficient rows compare five weighting schemes, visually grouped as four stand-alone indicators and three coefficients from education-based relational specifications. All point estimates are negative. Mutually adjusted parental rank has intervals spanning zero under DBS and selection weights, while its unit-weight interval is below zero.

## 4 Discussion

Socioeconomic patterning of KDM age acceleration appeared across both stand-alone SES indicators and education-based relational specifications, with gradient magnitude differing across SES representations. All four stand-alone indicators were inversely associated with KDM age acceleration, but the associations were stronger for wealth and income than for respondent and parental education; differences within the two education indicators and within the two material-resource indicators were small. The relational specifications showed inverse coefficients for educational mobility and mutually adjusted respondent rank, whereas the mutually adjusted parental-rank coefficient was close to zero. Across countries, direction was broadly shared while between-country heterogeneity varied across SES representations. Higher acceleration also predicted functional limitations, poorer cognition and mortality. Together, these findings demonstrate that socioeconomic patterning of biological ageing varies both with how socioeconomic position is represented and with the national setting in which it is observed.

The differences across SES representations suggest that socioeconomic patterning of biological ageing is not well described by a single socioeconomic gradient. The stronger gradients for wealth and income may reflect the proximity of material resources to later-life living conditions and physiological exposures, although current economic position can also incorporate health selection and reverse causation. The relatively small differences within the education pair and within the material-resource pair further suggest that the largest contrasts were between forms of socioeconomic position rather than between closely related indicators. The inverse associations for educational mobility and respondent attainment conditional on parental background are consistent with adult socioeconomic attainment remaining relevant when social origin is taken into account, whereas the near-zero conditional parental-rank coefficient indicates little additional association of parental educational rank once respondent rank is represented in the same model. This coefficient should be interpreted in light of the coarse retrospective measurement of parental education and the possibility that adult attainment embodies pathways originating earlier in life [12].

The cross-national results add a distinct comparative dimension. Predominantly inverse country-specific estimates point to broad commonality in the direction of socioeconomic patterning, while the extent of heterogeneity differed across SES representations. This combination suggests that socioeconomic disadvantage may be physiologically patterned across diverse national settings without implying a uniform gradient magnitude, extending European evidence on mortality and healthy-ageing inequalities to a multisystem physiological measure [20–22]. Such variation raises the question of whether national institutions modify the strength of socioeconomic gradients: welfare provision, pension protection, healthcare financing and labour-market conditions may alter both exposure to disadvantage and the resources available to buffer its consequences [17]. The exploratory country-context analyses did not reveal a consistent pattern across the examined contextual indicators, but the small number of countries provided limited information for distinguishing among competing institutional explanations [43]. Larger comparative samples with historically aligned contextual measures may be better suited to identifying the conditions underlying representation-specific variation [44].

The prospective associations with functional limitations, cognition and mortality support the health relevance of the physiological variation captured by the KDM measure, consistent with previous evaluations of KDM-based biological age [23–26, 32]. The recurrence of the main socioeconomic pattern under alternative weighting schemes and with the reduced biomarker panel further indicates that the findings were not tied to a single weighting strategy or biomarker composition. Similarity between official DBS-weighted and SIPW estimates supports stability to reweighting for measured predictors of complete-panel availability [45], while the reduced-panel analyses show that the overall pattern persisted when biomarker eligibility was broadened. Strengths of the study include the examination of multiple representations of socioeconomic position within a common survey and biomarker framework across 12 countries, explicit estimation of between-country heterogeneity, multiple-imputation uncertainty for monetary measures and fixed within-country reference scales, which kept SES units stable across analyses and defined coefficients as gradients in relative socioeconomic position within national distributions.

Several limitations qualify these findings. The socioeconomic analyses were cross-sectional and therefore associational, with possible confounding, health selection and reverse causation, especially for current economic resources. Complete-indicator requirements, selective survival, medication use and residual DBS field-condition variation may also affect estimates [46, 47]. Uneven country sample sizes and weight dispersion limited precision, and respondent-level PSU substitution approximated unavailable sampling clusters. Reported uncertainty conditions on estimated KDM calibration and SES reference scales. Longitudinal biomarker measurements and richer socioeconomic histories could establish when these gradients in physiological ageing emerge and how they evolve over the life course, while larger comparative datasets with historically harmonised institutional measures could clarify why some socioeconomic gradients are more stable across countries than others and which social and policy environments contribute to that variation.

## 5 Conclusion

Socioeconomic gradients in KDM age acceleration were predominantly inverse but varied in magnitude across representations of socioeconomic position. Their direction was broadly shared across 12 European countries, while the extent of between-country heterogeneity differed across representations. These findings support a multidimensional and comparative view of socioeconomic patterning in biological ageing, in which both the representation of socioeconomic position and the national setting shape the observed gradient.

## Data Availability

The SHARE data analyzed in this study are available for scientific use to registered researchers through the SHARE Research Data Center, subject to the SHARE Conditions of Use.

http://share-eric.eu/data/data-access

## Ethics approval

SHARE Waves 6–9 were reviewed and approved by the Ethics Council of the Max Planck Society, with additional country-specific review where required [48]. Necessary national permissions and ethics approvals preceded Wave 6 DBS fieldwork, and written informed consent was required for blood collection and future analysis [30, 31]. This study was a secondary analysis of released SHARE data and involved no new specimen collection.

## Supplementary data

Supplementary methods, results and Tables S1–S8 accompany this article.

## Data availability

The data used in this study are available to registered users from the Survey of Health, Ageing and Retirement in Europe (SHARE), subject to SHARE’s conditions of use. The analyses used SHARE Wave 6 interview and generated-variable data (Release 9.0.0), SHARE dried-blood-spot biomarker data (Release 2.0.0), Gateway Harmonized SHARE Version G, and SHARE Waves 7–9 (Release 9.0.0). The analytic code is provided in the Supplementary Material.

## Use of artificial intelligence (AI) tools

ChatGPT was used to improve the readability of the manuscript. The author takes full responsibility for the content of the manuscript.

