## appendix for "Multidimensional socioeconomic gradients in biological age acceleration across 12 countries in the SHARE study" (from another version of this preprint: version 1, 2026-09-16)

### Supplementary Appendix

Socioeconomic inequalities in biological age: multidimensional gradients and cross-national heterogeneity in SHARE

#### A Supplementary methods and analyses

This appendix provides technical details and complete results for analyses summarized in the main text. It covers survey design and KDM construction; monetary variables and multiple imputation; prospective predictive validity and measurement properties; pooled and country-varying socioeconomic models; sex and baseline-age heterogeneity; exploratory country-context moderation; descriptive country profiles; and weighting and selection sensitivity. Omnibus tests with Benjamini–Hochberg correction provide the inferential summaries, whereas pointwise margins and country-profile values are descriptive.

##### A.1 Survey-design and KDM construction details

###### A.1.1 Analytic survey design

All primary analyses used the official Wave 6 DBS analysis weight. Released strata and primary sampling unit (PSU) identifiers were nested within country and released SHARE subsample. Analytic strata cross-classified country, subsample, and the two released stratification fields. For records with a nonblank released PSU in a country-by-subsample domain containing at least two distinct nonblank released PSUs, the analytic PSU nested country, subsample, and released PSU; all other records received a unique respondent-level analytic PSU within the domain. This respondent-level fallback was a study-specific rule for incomplete or insufficient released PSU identifiers and did not reconstruct unobserved original sampling clusters. Survey designs were specified with `ids = ~analysis_psu`, `strata = ~analysis_strata`, `weights = ~analysis_weight`, and `nest = TRUE`; lonely-PSU strata used the `survey` adjustment rule. Design-based estimation was implemented with the `survey` package [1].

Descriptive statistics were calculated separately for the three panel-specific samples using the official DBS weight. Continuous variables were summarized by weighted means and standard deviations and categorical variables by weighted proportions. Kish effective sample size was reported as a diagnostic of weight dispersion [2]. The KDM9, KDM7, and KDM5 samples of 10,043, 10,581, and 14,015 participants corresponded to Kish effective sample sizes of 1,186, 1,267, and 1,867, respectively. Wealth and income ranks were averaged across the five released multiply imputed monetary datasets (SHARE "implicates") for descriptive presentation but retained separately by implicate in inferential models.

##### A.1.2 KDM estimation and age acceleration

KDM parameters were estimated separately by sex and biomarker panel using the official DBS weight. For biomarker $j$, I fitted the weighted biomarker-on-age regression

$$X_{ij} = q_j + k_j A_i + \varepsilon_{ij},$$

where $X_{ij}$ is the biomarker value for respondent $i$, $A_i$ is chronological age, $q_j$ and $k_j$ are the fitted intercept and age slope, and $s_j$ is the weighted residual standard deviation. The KDM biological-age estimate was

$$\mathrm{KDMBA}_i = \frac{\sum_{j=1}^{m}(X_{ij} - q_j)k_j/s_j^2 + A_i/s_{BA}^2}{\sum_{j=1}^{m}(k_j/s_j)^2 + 1/s_{BA}^2},$$

where $m$ is the number of biomarkers. Let $r_j$ denote the squared official-weighted correlation between biomarker $j$ and age. The implementation used the following finite-age-range correction:

$$r_{\mathrm{char}} = \frac{\sum_j |(k_j/s_j)\sqrt{r_j}|}{\sum_j |k_j/s_j|}, \qquad s_r = \frac{1 - r_{\mathrm{char}}^2}{r_{\mathrm{char}}^2}\frac{(75-50)^2}{12m}.$$

Biomarker-implied age was

$$A_i^M = \frac{\sum_j (X_{ij} - q_j)k_j/s_j^2}{\sum_j (k_j/s_j)^2},$$

and $s_{BA}^2$ was the official-weighted variance of $A_i^M - A_i$ minus $s_r$. At least 500 complete observations per sex were required to estimate a panel's biomarker–age parameters, and each KDM score required complete data on all biomarkers in its panel.

Raw KDM biological age was converted to age acceleration separately by sex. I regressed raw KDM biological age on a natural cubic spline of chronological age with three degrees of freedom using the official DBS weight and divided the residual by its sex-specific weighted standard deviation. The resulting KDM9, KDM7, and KDM5 acceleration scores had weighted means of approximately zero and weighted standard deviations of one within sex.

#### A.2 Socioeconomic variables, ranks, and multiple imputation

##### A.2.1 Education and educational ranks

Respondent education used reported years of schooling, retaining values from 0 to 25. Maternal and paternal ISCED-97 categories 0–6 were mapped to 0, 6, 9, 12, 13, 16, and 17 years, respectively. Highest-parent education was the maximum available mapped value; when only one parent's education was available, that value was used. Respondent and highest-parent education were each standardized using their official-weighted country-specific mean and standard deviation among eligible Wave 6 biomarker respondents with the corresponding variable available.

Educational origin and destination ranks used highest-parent and respondent education, respectively. Within each country and birth-cohort band, official-weighted midranks were calculated among eligible Wave 6 biomarker respondents with both measures available, using the same midrank formula given below for monetary variables. The birth-cohort bands were 1929 or earlier, 1930–1939, 1940–1949, 1950–1959, and 1960 or later. Origin and destination ranks were divided by 10 for modeling, so their coefficients represent a 10-percentile-point increase within country and birth-cohort band. Residualized educational-rank mobility was obtained from an official-weighted regression of destination rank on origin rank with an intercept fitted separately in each country and cohort band; the resulting residuals were standardized using their official-weighted mean and standard deviation within country. These reference samples were defined before applying the complete-marker restrictions of the individual KDM panels.

##### A.2.2 Wealth and income construction

Total wealth was reconstructed separately in each of the five released Wave 6 monetary implicates as

$$W = home + ores + car + vbus(sbus/100) + slti + bsmf + bacc - liab - mort,$$

and was retained only when the released Wave 6 asset-total gateway indicated a valid total. Individual income was the exact sum of `ydip`, `yind`, five `ypen` items, five `ylsp` items, `yreg1`, `yreg2`, `ylsr1`, and `ylsr2`. Couple income summed individual income across partners within the released couple identifier. Relative household income added `ysrent` and `ybabsmf` to couple income and was retained only when the released income-total gateway indicated a valid total. The reconstructed measures reconciled to the released totals in implicate 1.

Within each country and implicate, wealth and income were converted to official-weighted midranks. For value $x_j$,

$$P_j = 100 \frac{\sum_{k:x_k<x_j} w_k + \frac{1}{2} \sum_{k:x_k=x_j} w_k}{\sum_k w_k}.$$

The reference population comprised positive-weight respondents in the eligible Wave 6 biomarker sample in the relevant country and implicate whose construct was available in all five implicates. Percentile ranks were divided by 10 for modeling.

##### A.2.3 Released monetary implicates

I used the five released Wave 6 monetary implicates (`implicat`=1,...,5). SHARE monetary imputation uses fully conditional specification with iterative Gibbs sampling [3, 4]. Monetary variables are generally modeled with a two-part procedure comprising an ownership model and a conditional amount model, with bounds from unfolding brackets or component information and transformations for skewed amounts. Hot-deck or regression fallback may be used when donor support is insufficient. Models are generated separately by country and household type.

Within this study, wealth and income were reconstructed and ranked separately in every implicate. Respondents were retained only when the construct was available in all five implicates, and identical respondent IDs, outcomes, survey-design variables, covariates, and country sets were used in each implicate-specific model. Estimates were combined using Rubin's rules [5]. For scalar estimate $Q_m$ with within-implicate variance $U_m$,

$$\bar{Q} = \frac{1}{M}\sum_{m=1}^{M} Q_m, \qquad \bar{U} = \frac{1}{M}\sum_{m=1}^{M} U_m, \qquad B = \frac{1}{M-1}\sum_{m=1}^{M}(Q_m - \bar{Q})^2,$$

$$T = \bar{U} + \left(1 + \frac{1}{M}\right) B,$$

with $M = 5$. Scalar confidence intervals and tests used $\sqrt{T}$ with finite-complete-data Barnard–Rubin degrees of freedom [6]. Multivariate hypotheses and country-slope contrasts used the corresponding pooled coefficient vectors and within-, between-, and total-covariance matrices, retaining both sampling and between-implicate uncertainty.

#### A.3 Prospective predictive validity and measurement properties

##### A.3.1 Outcome samples and models

Each KDM measure was evaluated in its own panel-specific outcome-eligible sample. Incident limitation on the six-item ADL and nine-item IADL measures at Wave 9 was the main functional endpoint; no limitation at Wave 6 and any limitation at follow-up defined incidence. Corresponding outcomes at Waves 7 and 8, the Wave 9 four-domain cognitive composite, and mortality through follow-up provided complementary predictive-validity evidence. Functional outcomes were modeled with survey-weighted quasibinomial regression with a logit link, multidomain cognition with survey-weighted Gaussian regression adjusted for the same Wave 6 composite, and mortality with survey-weighted Cox regression. All models retained the demographic adjustment set and SHARE survey design described in the main Methods.

An outcome-model cell was reported only when it included at least 500 respondents, had a Kish effective sample size of at least 250, and retained at least 10 survey-design degrees of freedom. Binary and mortality cells additionally required at least 50 events. Observed mortality follow-up was summarized without survey weights: median follow-up was 6.67 years (IQR, 5.67–6.83) for KDM9, 6.58 years (IQR, 5.12–6.83) for KDM7, and 6.67 years (IQR, 5.00–6.83) for KDM5; the corresponding death counts were 560, 623, and 811.

For mortality, released year–month dates were represented by month midpoints; a recorded year with an unavailable month retained the midyear convention. A valid death date within seven years defined an event at that time, and a valid later death date established survival through the seven-year administrative horizon. Without a valid death date, censoring occurred at the last regular Main interview in Waves 6–9, capped at seven years. End-of-life proxy contacts established death-status information but their interview dates did not establish survival time. Records with unresolved death dates were explicitly flagged, and records without positive follow-up were excluded from Cox estimation. Raw dates flagged as implausible remained invalid when source review could not establish a credible replacement. Undated deaths therefore retain unresolved event times.

Table S1 reports the complete panel-specific predictive associations, eligible sample sizes, event counts, effect scales, confidence intervals, and unadjusted $P$ values. For the 18 panel-specific functional estimates, all 18 confidence intervals excluded 1. Figure S1 displays the earlier-wave functional results that complement the Wave 9 functional, cognitive, and mortality results shown in the main predictive-validity figure.

##### A.3.2 Cognitive composite construction

The calibration sample comprised eligible Wave 6 biomarker respondents with positive official DBS weights and complete immediate recall, delayed recall, animal fluency, serial-seven numeracy, and temporal orientation scores at Wave 6. Calibration was performed separately within each country on this baseline sample. Each task was first standardized using its official-weighted Wave 6 mean and standard deviation. The immediate- and delayed-recall task z scores were averaged, and this average was then restandardized using its own country-specific weighted Wave 6 mean and standard deviation to form the episodic-memory domain. The memory-domain z score and the three remaining task z scores were averaged with equal weights. This four-domain average was finally standardized using its country-specific weighted Wave 6 mean and standard deviation. The same baseline parameters at all three stages—task, memory-domain, and final-composite standardization—were applied unchanged to Wave 9. Scores required all five tasks at the relevant wave and were expressed in within-country Wave 6 cognitive SD units. Baseline calibration preceded KDM-panel and follow-up-availability restrictions.

##### A.3.3 Overlap among KDM measures

Table S2 reports pairwise Pearson correlations among the age-residualized, within-sex standardized KDM scores with and without the official DBS weight. Each pair uses all respondents with both scores observed, without additional SES restrictions; the table note also reports the common three-score sample check. The correlations quantify the degree of overlap among the KDM panels.

#### A.4 Primary pooled and country-varying model details

Let $Y_i$ denote a KDM acceleration outcome, $C_i$ country, $f_3(A_i)$ a natural cubic spline of Wave 6 age with three degrees of freedom, $S_i$ sex, and $g_s(X_i)$ the exposure term for SES specification $s$. The common-slope model was

$$\text{M0}: \quad Y_i = \alpha + g_s(X_i) + \sum_{c \neq c_0} \gamma_c I(C_i = c) + f_3(A_i) + \delta S_i + \varepsilon_i,$$

and the country-varying model was

$$\text{M1}: \quad Y_i = \alpha + g_s(X_i) + \sum_{c \neq c_0} \gamma_c I(C_i = c) + \sum_{c \neq c_0} \eta_{cs} g_s(X_i) I(C_i = c) + f_3(A_i) + \delta S_i + \varepsilon_i.$$

Germany served as the reference country in every model in which it was included. For respondent education, highest-parent education, wealth rank, income rank, and residualized educational-rank mobility, $g_s(X_i)$ comprised one continuous exposure. The joint origin–destination model included separate $R^O/10$ and $R^D/10$ terms and, in M1, separate country interactions for both. Its pooled specification-level hypothesis therefore had two degrees of freedom; the other five specifications had one.

Country-specific slopes were obtained as linear combinations of coefficients from the single pooled M1 fit. Country heterogeneity was tested by jointly constraining all nonreference-country exposure interactions to zero; for the joint origin–destination specification, both sets of interactions were included. Within each KDM outcome, the Benjamini–Hochberg procedure [7] was applied across the six pooled specification-level tests and, separately, across the six country-heterogeneity tests.

#### A.5 Complete estimates for the socioeconomic analysis

Table S3 provides the complete pooled coefficient set underlying the primary socioeconomic analysis. For each KDM outcome, the table includes five single-coefficient SES specifications and the two conditional coefficients from the joint parental-origin–respondent-destination rank model, for 21 displayed coefficients in total. The joint origin–destination specification is treated as one of the six SES specifications and is evaluated with a two-degree-of-freedom block test; its adjusted $P$ value therefore applies to the joint block and is shared by the two displayed component rows.

Across the three KDM measures, pooled coefficients ranged from −0.107 to −0.076 SD per 1-SD higher respondent education, from −0.075 to −0.066 SD per 1-SD higher highest-parent education, from −0.076 to −0.061 SD per 10-percentile higher wealth rank, and from −0.074 to −0.060 SD per 10-percentile higher income rank. Under the fitted linear rank models, a contrast from the 10th to the 90th percentile corresponded to 0.49–0.61 SD lower KDM age acceleration for wealth and 0.48–0.59 SD lower KDM age acceleration for income.

Association and SES-by-country heterogeneity tests were adjusted in separate six-test BH families within each

KDM outcome. All 18 pooled specification tests had BH-adjusted $P < 0.05$, as did 11 of the 18 omnibus SES-by-country heterogeneity tests.

##### A.5.1 Magnitude of country heterogeneity

For each of the 21 specific SES–KDM contrasts, Table S4 reports the pooled coefficient, minimum and maximum estimated country slope, number of negative point estimates, number of pointwise 95% confidence intervals wholly below zero, and estimated between-country standard deviation $\tau$. The 18 omnibus specifications contain 21 contrasts because each joint origin–destination model supplies two coefficients. Summaries use all countries included for each contrast: 12 for respondent education, wealth, and income, and 11 for highest-parent education, residualized educational-rank mobility, and the joint origin–destination specification. Counts describe point estimates or pointwise intervals and do not define a practical-equivalence threshold.

Let $\hat{\boldsymbol{b}}$ denote the country-slope vector and $V_{\text{full}}$ its full sampling covariance, including Rubin's total covariance for monetary exposures. For a one-contrast specification, the moderator-free model is

$$\hat{\boldsymbol{b}} = \mu\mathbf{1} + \boldsymbol{u} + \boldsymbol{e}, \qquad \boldsymbol{u} \sim N(\mathbf{0}, \tau^2 I), \qquad \boldsymbol{e} \sim N(\mathbf{0}, V_{\text{full}}).$$

Variance components are estimated by restricted maximum likelihood. Joint origin–destination models use contrast-specific intercepts and diagonal contrast-specific random-effect variances, retaining the full sampling covariance between countries and contrasts. The displayed $\tau$ is the square root of the moderator-free variance; it quantifies total between-country dispersion before adding the country-context indicators described below. Its unit is KDM acceleration SD per displayed SES increment. The coefficient range contains sampling variation as well as between-country differences, while $\tau$ estimates the latter under the random-effects model. These variance-component estimates carry uncertainty with only 11 or 12 countries; estimates near the boundary do not establish exact homogeneity. The table presents point estimates of $\tau$, with no small/moderate/large categorization. For KDM9, respondent-education slopes ranged from $-0.213$ to 0.054 SD and income-rank slopes from $-0.115$ to 0.030 SD per 10-percentile increment; 11 of 12 point estimates were negative for each exposure, and the corresponding estimates of $\tau$ were 0.072 and 0.022. The complete table provides the remaining contrasts.

#### A.6 Sex and baseline-age heterogeneity

##### A.6.1 Model construction and multiplicity

Secondary heterogeneity analyses crossed three KDM outcomes with six SES specifications and three interaction models. The sex model added SES-by-sex terms. The linear-age model added SES-by-linear-age terms while retaining the three-degree-of-freedom natural cubic spline for the age main effect. A nonlinear sensitivity model interacted each SES term with all three age-spline basis functions. The joint origin–destination specification was tested as a multiparameter block. Because all baseline measurements were obtained in the same survey wave, age contrasts cannot be separated from birth-cohort differences and may also be affected by selective survival.

All models retained the official SHARE DBS weight, complex-survey design, and the SES-by-country terms from the country-varying association model. The additional SES-by-sex and SES-by-age coefficients were common across countries. Wealth and income models were fitted separately in the five released monetary implicates. Benjamini–Hochberg correction was applied across the six SES specifications within each KDM outcome and interaction model [BH(6)]. Within each outcome, the six sex and six linear-age tests were also adjusted jointly as a 12-test family [BH(12)]. The nonlinear spline tests formed a separate sensitivity family.

##### A.6.2 Global interaction evidence

All 54 interaction tests were estimable. Two sex interactions had unadjusted $P < 0.05$, both for KDM7: highest-parent education, $F(1, 6454) = 4.65$, $P = 0.031$, and respondent education, $F(1, 6938) = 4.51$, $P = 0.034$. Neither had a BH-adjusted $P < 0.05$ under the six- or 12-test adjustment. No linear-age or nonlinear age-spline interaction had a BH-adjusted $P < 0.05$; the smallest unadjusted linear-age $P$ value was 0.177, and the smallest nonlinear spline $P$ value was 0.211.

The two nominal KDM7 sex interactions corresponded to more negative education coefficients among women at age 62.5 years. For respondent education, the estimated association was −0.158 SD (95% CI, −0.239 to −0.077) among women and −0.054 SD (95% CI, −0.112 to 0.004) among men. For highest-parent education, the corresponding estimates were −0.125 SD (95% CI, −0.206 to −0.043) and −0.008 SD (95% CI, −0.076 to 0.060).

Figure S2 and Table S5 report the complete omnibus tests. Figure S3 and Table S6 provide descriptive KDM9 margins. The omnibus tests assess systematic modification; the pointwise margins illustrate the fitted interaction surfaces.

#### A.7 Exploratory country-context moderation and covariance sensitivity

##### A.7.1 Contextual measures and second-stage model

Table S13 provides the source locators, extraction date, and selection and transformation rules for the six moderators, including the sex averaging of pension replacement rates and the definition selection for OECD Gini records.

The six contextual measures characterized the Wave 6/DBS policy and economic environment. Public social expenditure was the 2015 percentage of GDP from the OECD Social Expenditure Database [8]. Disposable-income inequality was the 2015 after-tax-and-transfer Gini coefficient from the OECD Income Distribution Database [9]. The low-earner pension measure was the net pension replacement rate at 50% of average earnings under 2014 policy rules from *Pensions at a Glance 2015* [10]. Out-of-pocket health expenditure was the 2015 percentage of current health expenditure from the WHO Global Health Expenditure Database, accessed through World Development Indicators [11, 12]. Economic development was the natural logarithm of 2015 GDP per capita at purchasing-power parity in constant 2021 international dollars, and unemployment was the 2015 ILO modeled estimate as a percentage of the total labor force, both from World Development Indicators [12]. Near-baseline measures were used to maximize consistency of definition and coverage across the participating countries. They were treated as parsimonious indicators of national context rather than as direct reconstructions of cumulative life-course institutional exposure; assembling multi-decade trajectories would require additional historical harmonization and explicit assumptions about exposure windows, accumulation, and institutional discontinuities.

Each moderator was standardized with equal country weights across the 12 DBS countries. All six moderators were examined for each of the 18 outcome–SES combinations, giving 108 models; inclusion of a combination was independent of its country-heterogeneity test result. Each model used the 11 or 12 countries represented in its first-stage SES specification. For an outcome–SES cell, let $\widehat{\boldsymbol{\beta}}$ and $\mathbf{T}_\beta$ denote the Rubin-pooled M1 coefficient vector and total covariance matrix. A contrast matrix $\mathbf{L}$ extracted all country-specific slopes included in that analysis:

$$\widehat{\mathbf{b}} = \mathbf{L}\widehat{\boldsymbol{\beta}}, \qquad \mathbf{V}_{\text{full}} = \mathbf{L}\mathbf{T}_\beta\mathbf{L}'.$$

The full sampling covariance retained covariance induced by the common M1 fit, including off-country covariance and, for the joint origin–destination model, covariance between origin and destination contrasts.

For a single-exposure SES specification, the second-stage model was

$$\widehat{b}_c = \alpha + \theta z_c + u_c + e_c, \qquad u_c \sim N(0, \tau^2), \qquad \mathbf{e} \sim N(\mathbf{0}, \mathbf{V}_{\text{full}}),$$

fitted by multivariate restricted maximum likelihood with a country random intercept using `metafor` [13]. The joint origin–destination model used contrast-specific fixed intercepts, moderator coefficients, and random-effect variances while retaining the full sampling covariance.

Small-sample inference applied CR2 covariance adjustment clustered by country, Satterthwaite degrees of freedom for component tests, and an HTZ Wald test for the joint moderator hypothesis [14, 15]. Cluster wild-bootstrap inference used 1,999 Rademacher draws [16], with a plus-one Monte Carlo $P$ value [17]. These sources support the named procedures in their standard independent-cluster setting. Because $\mathbf{V}_{\text{full}}$ contained nonzero off-country blocks, applying country-cluster inference to the full-covariance model was treated as a study-specific exploratory extension and paired with the country-block-diagonal analysis described below. For each cell, I recorded the number of countries and unique moderator values, CR2 denominator degrees of freedom, maximum country leverage, completion of leave-one-country-out fits, and sign agreement across those fits. Potential instability was flagged for fewer than 10 countries or eight unique moderator values, CR2 denominator degrees of freedom of four or less, maximum country leverage of 0.50 or greater, incomplete leave-one-country-out fits, or less than 80% sign agreement. These indicators were reported descriptively rather than combined into an inferential pass/fail rule. Within each KDM outcome, BH correction was applied across the 36 SES-by-moderator tests separately for CR2 and wild-bootstrap inference.

##### A.7.2 Moderation results and diagnostics

Table S7 reports all 108 combinations of six SES specifications, six country-level moderators, and three KDM outcomes. No combination had a BH-adjusted $P < 0.05$ under either inferential method. Twenty-eight of 108 cells had no diagnostic flags; 69 were flagged for limited effective degrees of freedom, seven for high country leverage, two for inference-method sensitivity, and two for unstable leave-one-country-out direction. These diagnostics reinforce the exploratory status of the country-context analyses and limit their leverage for identifying institutional sources of heterogeneity. Figure S4 summarizes the omnibus results.

##### A.7.3 Paired covariance sensitivity

The paired sensitivity retained all variances and within-country covariances, including covariance between origin and destination contrasts, but set every off-country covariance element to zero. It therefore aligned the working sampling-covariance partition with the country units used for CR2, HTZ, and wild-bootstrap clustering. First-stage country slopes, included countries, moderators, REML specification, inferential engine, multiplicity family, and the same 1,999 Rademacher draws were otherwise held fixed.

Across 108 paired cells, the maximum absolute full-covariance versus block-diagonal differences in $P$ values were 0.047 for CR2 and 0.104 for CWB. No raw $P < 0.05$ classification changed for either method, and no CWB BH-adjusted decision changed. Because this was a same-engine paired sensitivity rather than an independent implementation, its decision concordance reduces evidence that the covariance choice changed the current null country-context conclusion but does not establish the theoretical validity of full-covariance country-cluster inference. The substantive summary retains the full-covariance results. Figure S5 and Table S8 provide the complete paired results.

#### A.8 Descriptive country association profiles

##### A.8.1 Complete-component set and profile construction

The country association-gradient summary provides a descriptive view of the country-specific coefficient pattern. Inclusion required all 21 country coefficients to be estimable:

$$(5 \text{ single-coefficient SES specifications} \times 3 \text{ KDM outcomes})$$
$$+ (2 \text{ origin–destination coefficients} \times 3 \text{ KDM outcomes}) = 21.$$

Eleven countries had all 21 coefficients available and entered the profile summary (Table S9). Greece retained 9 of 21 exact components and was excluded from the 11-country complete-profile display.

For each exact SES–outcome–coefficient component, the country-specific coefficient was multiplied by $-1$, so larger values denoted a stronger association of socioeconomic advantage with lower KDM acceleration, and standardized across the same 11 countries. For the joint origin–destination specification, standardized origin and destination values were first averaged within KDM outcome. The three KDM outcomes were then averaged with equal weight within each SES specification, yielding six country-standardized profile values per country. An equal-weight mean across the six displayed profile dimensions was then calculated for each country and used to order the visualization; no 0–100 rescaling was applied.

Table S10 reports the resulting SES-specific profile values. The country profiles visualize the multidimensional variation summarized by the omnibus country tests while making broad ordering patterns visible.

#### A.9 Survey- and selection-weight sensitivity

##### A.9.1 Target population and selection model

The selection analysis examined whether incomplete availability of the biomarker panels used to construct each KDM outcome materially altered the pooled SES associations. For KDM outcome $k$, $S_{ik} = 1$ indicated complete availability of the required biomarker panel. The selected samples contained 10,043 respondents for KDM9, 10,581 for KDM7, and 14,015 for KDM5, corresponding to official-weighted selected shares of 47.1%, 49.6%, and 72.1%.

The numerator model was intercept-only. The denominator model included country, sex, a three-degree-of-freedom natural spline of age, respondent education, highest-parent education, wealth decile, income decile, DBS batch, shipment time, drying time, recorded temperature, sample-opening and desiccant conditions, and a spline of log-transformed total blood-spot size [18, 19]. Missing highest-parent education, wealth decile, and income decile were replaced only within the selection model by country-specific medians with separate missingness indicators.

The denominator probability was estimated using official-weighted ridge logistic regression with an unpenalized intercept and $\lambda = 100$ for the remaining coefficients [20]. The stabilized multiplier and analysis weight were constructed from official DBS weight $w_i$, official-weighted selected proportion $\widehat{p}_N$, and fitted denominator probability $\widehat{p}_{Di}$, following general inverse-probability-weighting guidance [21, 22]:

$$r_i = \frac{\widehat{p}_N}{\widehat{p}_{Di}}, \qquad w_i^{\mathrm{SIPW}} = w_i r_i.$$

A trimmed SIPW analysis winsorized $r_i$ at the within-country 1st and 99th percentiles before applying it to the official weight. Predicted probabilities were numerically bounded to $[10^{-8}, 1 - 10^{-8}]$.

##### A.9.2 Weight diagnostics and point-estimate sensitivity

The official DBS-weighted primary models provided the reference for four alternative weighting schemes: within-country trimmed official DBS weights, unit weights, SIPW, and within-country P1/P99-trimmed SIPW. Unit weights assigned every included respondent a weight of one in the outcome regression. All schemes retained the existing KDM scores, SES transformations, and construction of the survey strata and PSU identifiers. Diagnostics included the maximum selected multiplier, effective sample size relative to the official-weight effective sample size, maximum absolute postweighting standardized mean difference, minimum denominator probability, and country coverage. At the point-estimation stage, a SIPW variant was included only when all five conditions were met: maximum multiplier no greater than 25, effective sample size at least 25% of the official-weight value, maximum absolute standardized mean difference no greater than 0.10, minimum denominator probability at least 0.01, and at least one selected respondent in every country of the eligible biomarker sample. The raw or trimmed multiplier and corresponding weights were used for the applicable variant. These joint diagnostic checks determined the SIPW variants included in the comparison.

Raw and trimmed SIPW met the joint point-stage conditions for KDM9 and KDM7. For KDM5, the raw SIPW maximum multiplier was 36.40, so this variant was excluded from the reported association comparison; within-country trimming reduced the maximum multiplier to 22.10, and the trimmed variant met all conditions and was included. Figure S6 and Table S11 present the pooled point-estimate comparisons.

Country inclusion was then assessed within each outcome–SES model under its applicable weights. For the continuous SES specifications, country support required at least 50 respondents, Kish effective sample size of at least 25, at least three distinct values for each exposure, and a weighted exposure standard deviation of at least 0.05 on the model scale. Joint origin–destination models additionally required an absolute weighted correlation below 0.98 between the two predictors. Monetary specifications required country support in every implicate. The same supported-country sample was used for the pooled and country-varying fits within a weighting scheme. Consequently, sample composition could differ between weighting schemes: for the KDM7 respondent-education model, Israel's 133 respondents had a country-specific effective sample size of 23.81 under raw SIPW and 23.84 under trimmed SIPW, below the threshold of 25. Both SIPW variants therefore used 10,448 respondents from 11 countries, compared with 10,581 respondents from 12 countries under the official weights.

##### A.9.3 Bootstrap propagation

Selection-model uncertainty was propagated using 1,000 Rao–Wu survey-bootstrap replicates [23]. The SIPW variants included at the point-estimation stage remained fixed throughout the bootstrap. Each replicate regenerated the survey multiplier, refitted the ridge selection model, reconstructed the applicable raw or trimmed SIPW, and refitted the outcome model subject to model-specific sample-support requirements. Replicate-specific weight diagnostics were retained without using them to reselect SIPW variants. A bootstrap-based uncertainty estimate was reported when at least 950 of the 1,000 replicates were successfully completed for that analysis unit. This completion requirement was separate from the point-stage diagnostic conditions used to include a weighting variant.

Bootstrap uncertainty estimates were available for all pooled sensitivity estimands across the SIPW analyses included in the point-estimate comparison. KDM7 also had complete availability for country-interaction coefficients and joint tests under raw and trimmed SIPW. For KDM9 under raw and trimmed SIPW and KDM5 under trimmed SIPW, most country-interaction coefficients were available but the corresponding joint country-interaction tests did not meet the bootstrap reporting criterion. Figure S7 and Table S12 report the complete availability results.

Taken together, the close agreement of the pooled estimates across official DBS, trimmed official DBS, SIPW, trimmed SIPW, and unit-weight analyses indicates that the pooled SES gradients were stable across the weighting schemes and associated sample-support restrictions examined here. Comparisons involving different country sets combine changes in weights and sample composition. This conclusion is strongest for the pooled associations; incomplete bootstrap availability for KDM9 and KDM5 country-interaction tests limits corresponding claims about selection robustness of the country-heterogeneity tests.

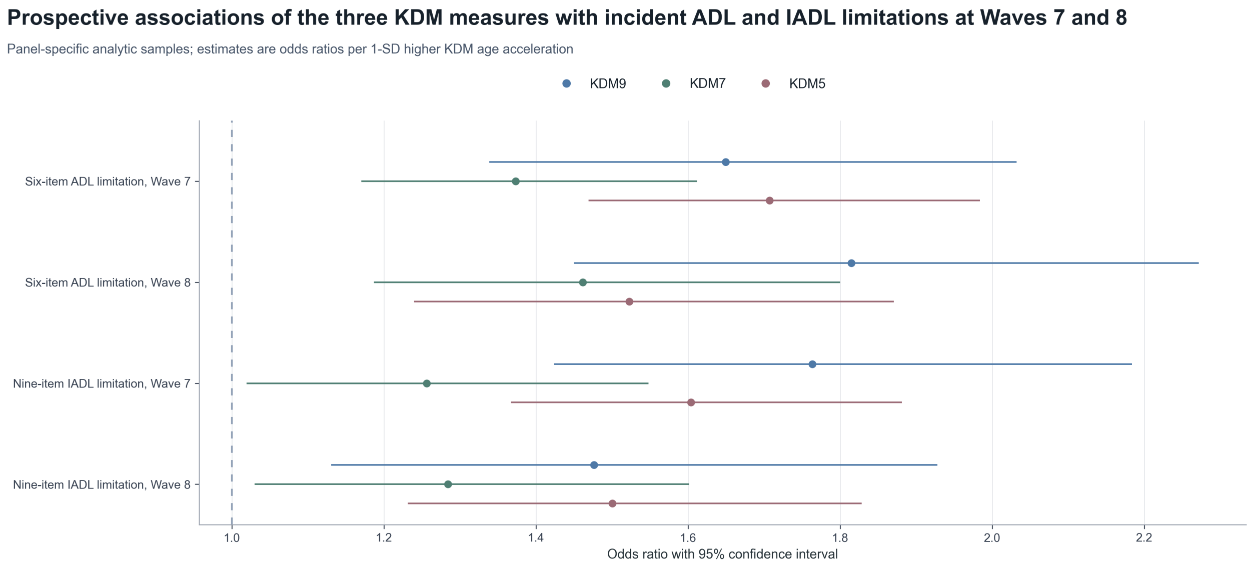


**Figure S1:** Prospective associations of the three KDM measures with incident limitation on the six-item ADL and nine-item IADL measures at Waves 7 and 8. Estimates use panel-specific samples and are odds ratios with 95% confidence intervals per 1-SD higher KDM age acceleration. These earlier-wave endpoints complement the Wave 9 functional endpoints in the main predictive-validity figure. Exact results are reported in Table S1.

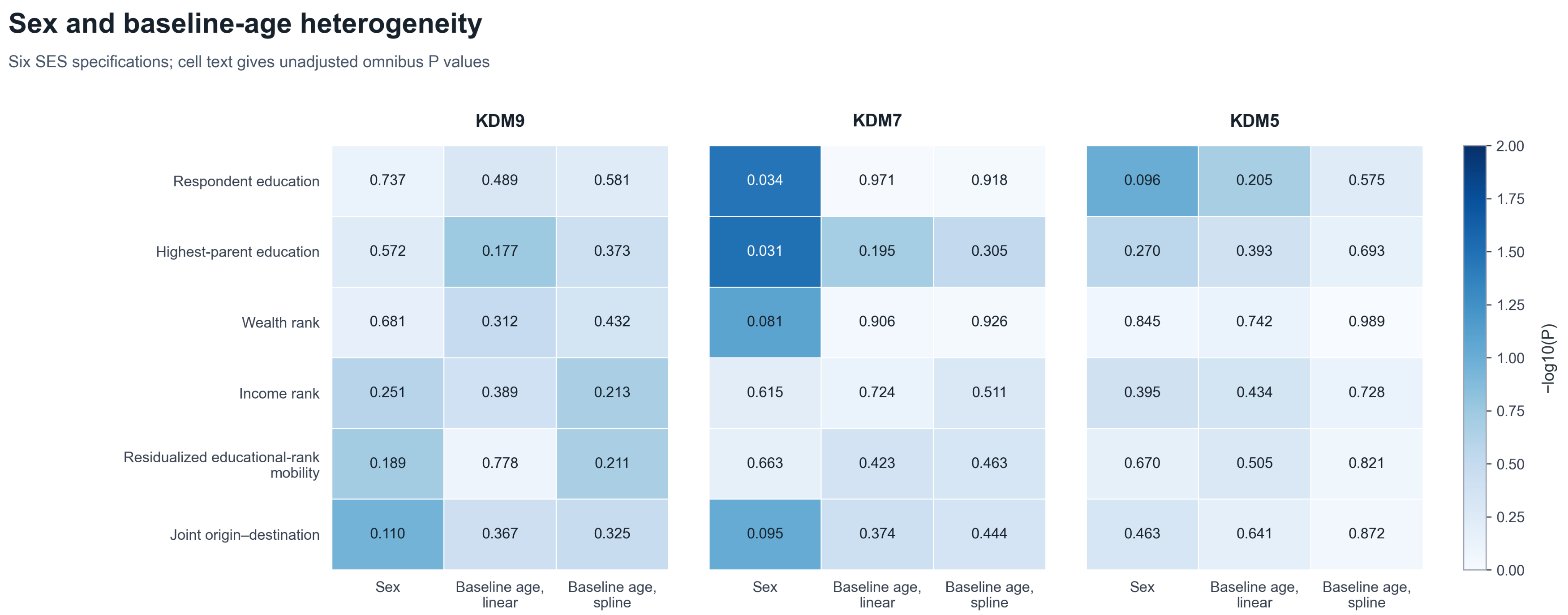


**Figure S2:** Omnibus tests of heterogeneity in socioeconomic associations by sex and baseline age across six SES specifications and three KDM outcomes. Cell text gives the unadjusted omnibus $P$ value and fill represents $-\log_{10}(P)$. None of the tests had a Benjamini–Hochberg-adjusted $P < 0.05$. Table S5 reports test statistics, degrees of freedom, and adjusted $P$ values.

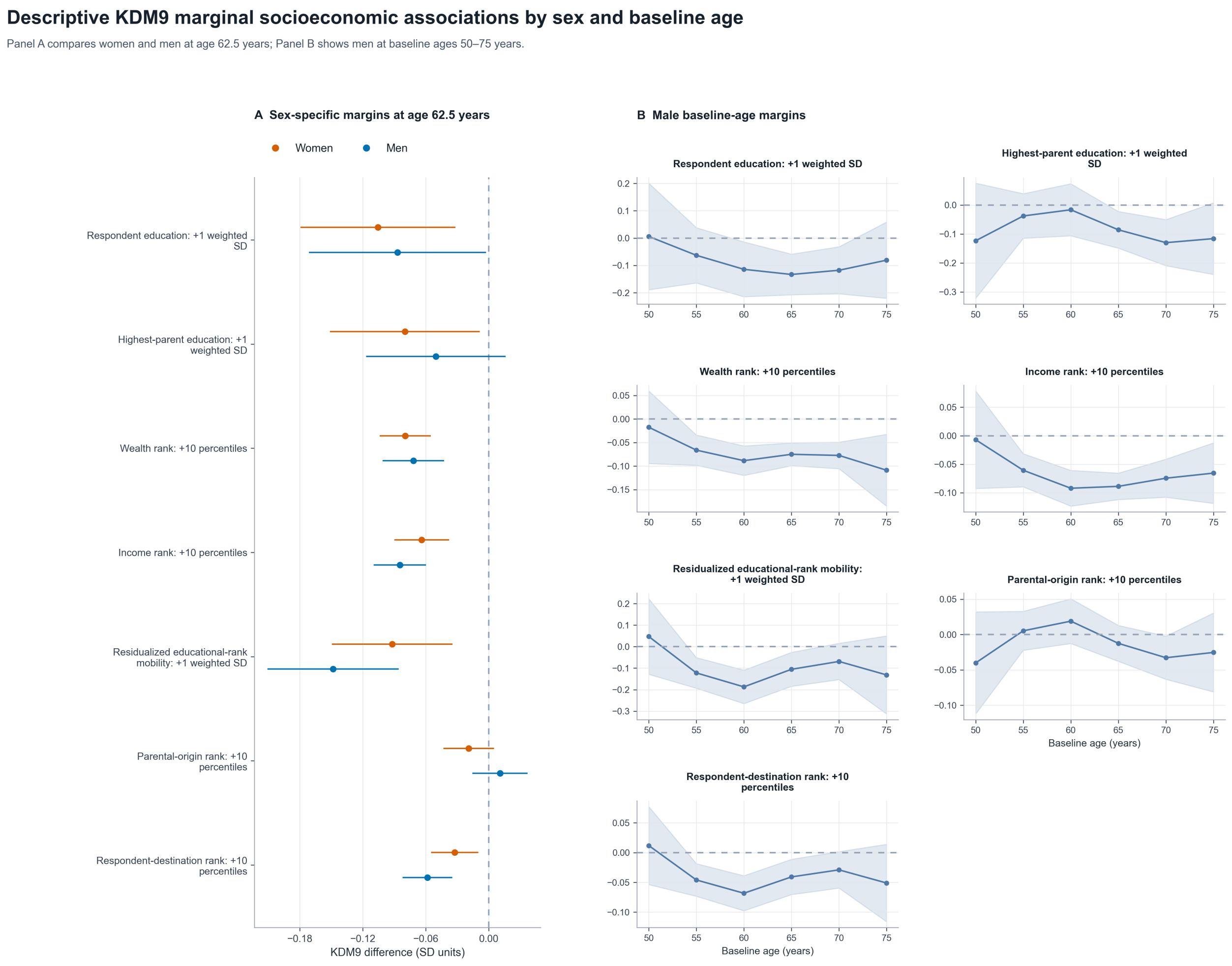


**Figure S3:** Descriptive KDM9 marginal socioeconomic associations by sex and baseline age. Panel A compares women and men at age 62.5 years. Panel B shows estimates for men at baseline ages 50–75 years. Intervals are pointwise 95% confidence intervals. Because all baseline measurements were obtained in the same survey wave, age contrasts cannot be separated from birth-cohort differences and may also be affected by selective survival. Global interaction tests are reported in Table S5, and exact margins are reported in Table S6.

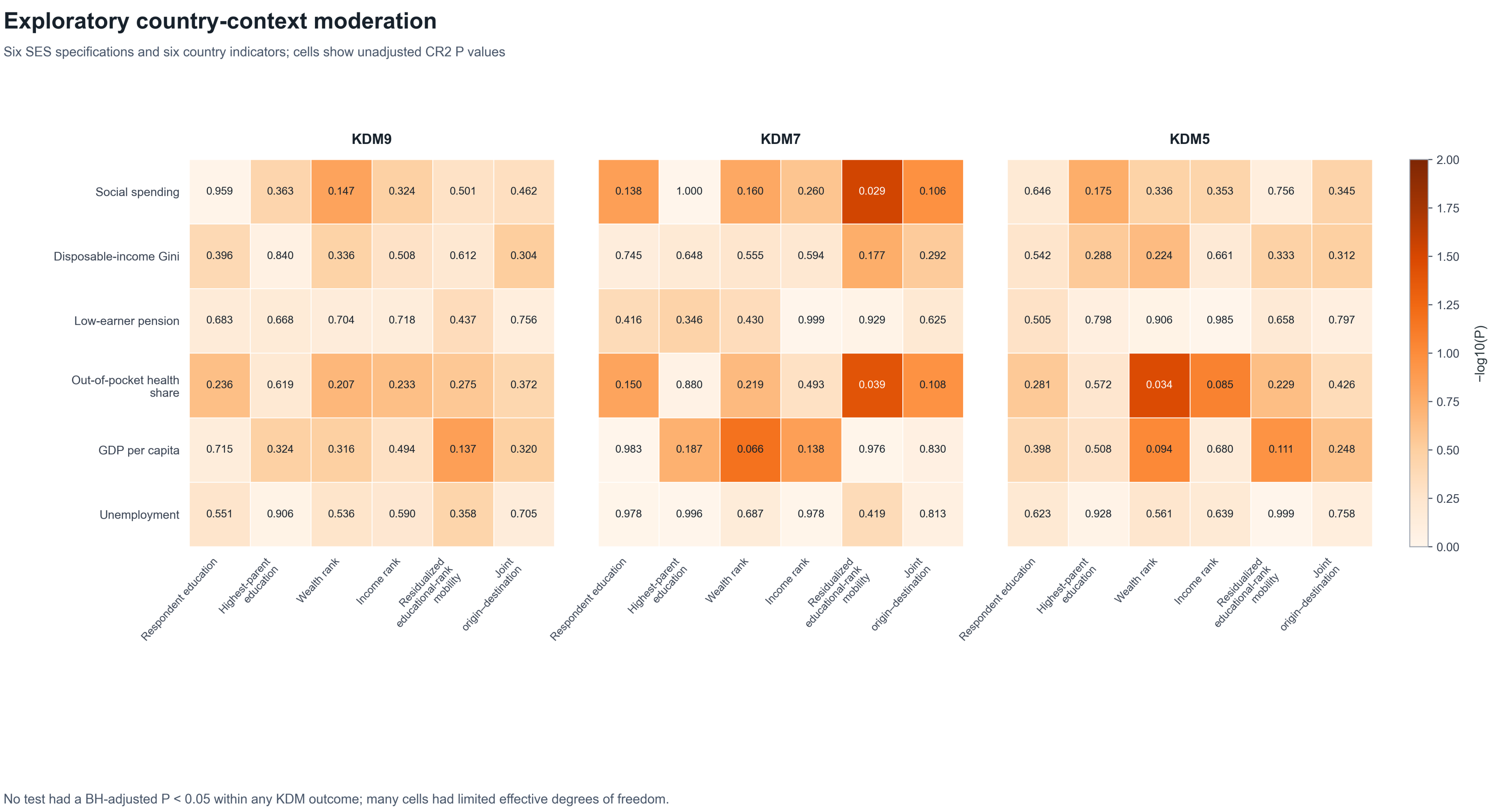


**Figure S4:** Exploratory country-context moderation across six SES specifications and three KDM outcomes. Cell text gives the unadjusted CR2 $P$ value and shading represents $-\log_{10}(P)$. Continuous CR2 and full-covariance country wild-bootstrap $P$ values are reported in Table S7. Neither method yielded a Benjamini–Hochberg-adjusted $P < 0.05$ for any outcome. Limited effective degrees of freedom constrained precision in many cells. These models assess simple relationships with contemporaneous national indicators rather than cumulative life-course institutional exposure; complete tests, adjusted $P$ values, and stability notes are reported in Table S7.

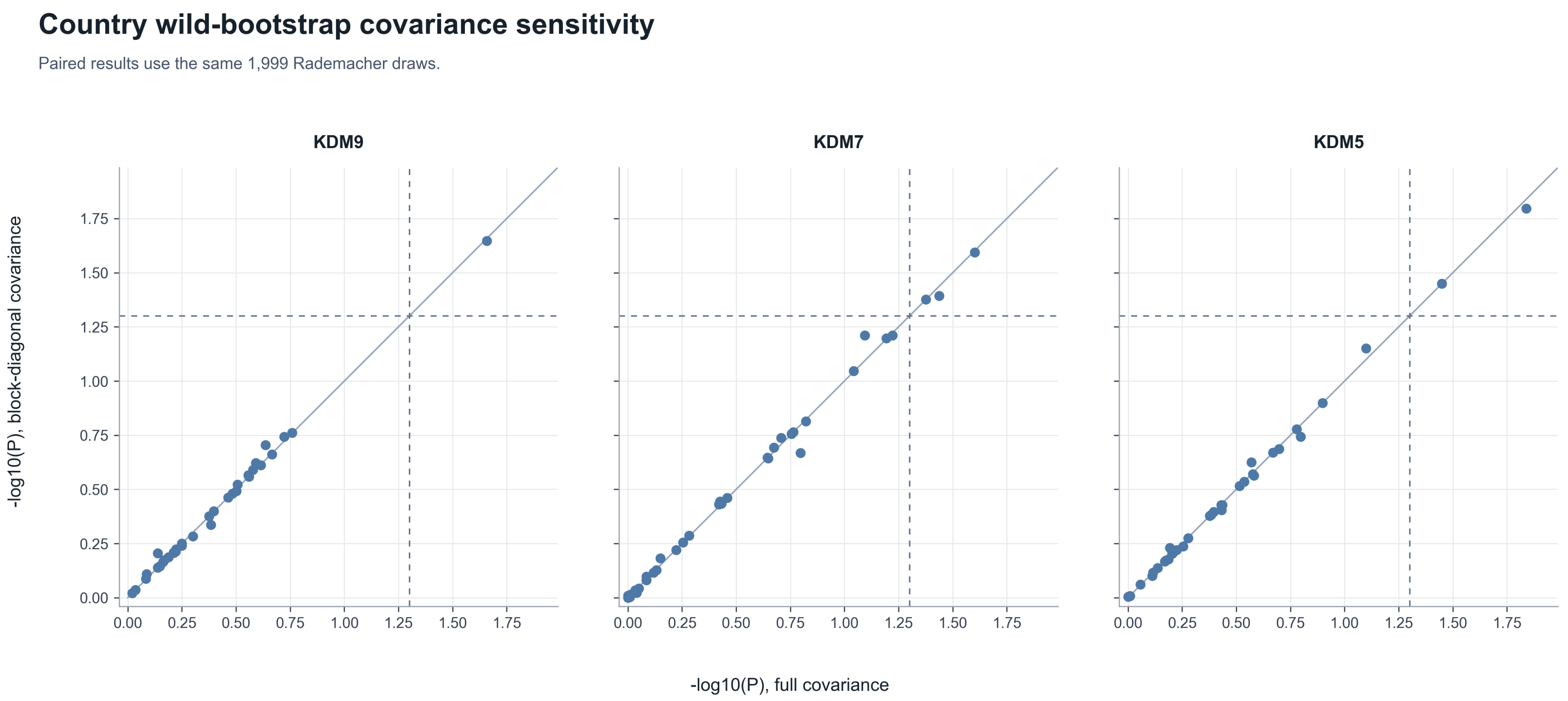


**Figure S5:** Paired country wild-bootstrap $P$ values under full country-slope covariance and block-diagonal covariance. Axes show $-\log_{10}(P)$; the diagonal denotes equality and dashed lines denote $P = 0.05$. Each pair uses identical country slopes, included countries, and the same 1,999 Rademacher draws. The two covariance specifications produced closely similar $P$ values, and BH-adjusted conclusions were unchanged; complete paired results are reported in Table S8.

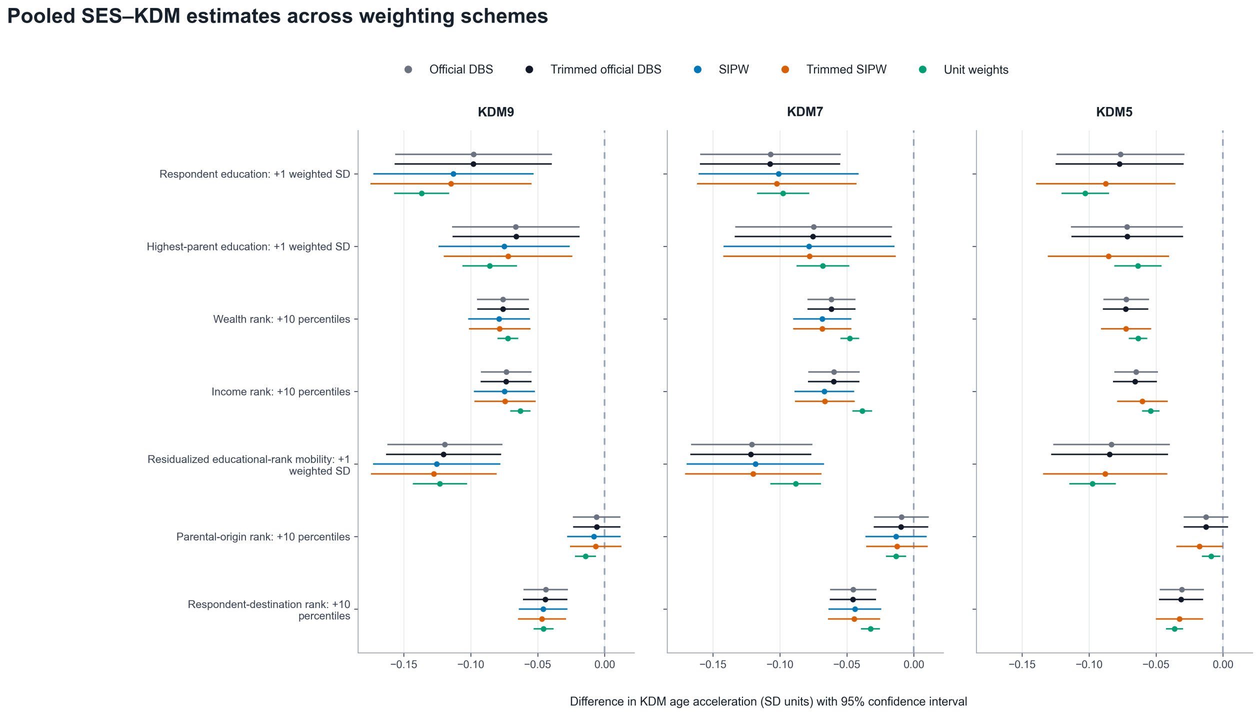


**Figure S6:** Pooled SES–KDM estimates across alternative weighting schemes. KDM9 and KDM7 include official DBS, trimmed official DBS, SIPW, trimmed SIPW, and unit-weight estimates; KDM5 omits SIPW because of greater weight instability (maximum multiplier, 36.40). Lines are 95% confidence intervals. Full point estimates and uncertainty are reported in Table S11.

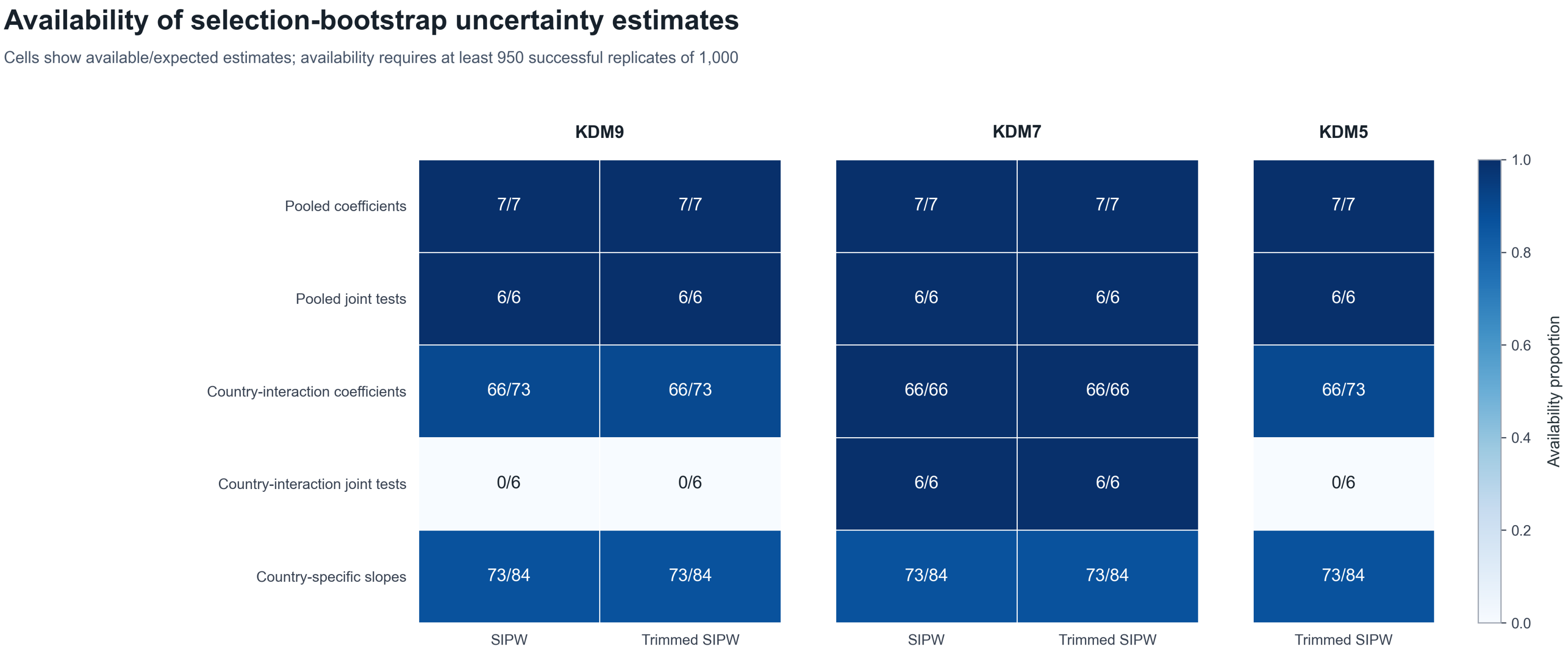


**Figure S7:** Availability of selection-bootstrap uncertainty estimates. Cell text gives the number of estimates available over the number expected, and fill represents the available fraction. A bootstrap uncertainty estimate was considered available when at least 950 of 1,000 replicates completed successfully. Complete availability results are reported in Table S12.

**Table S1:** Prospective associations of KDM9, KDM7, and KDM5 with ADL6, IADL9, and other age-related outcomes

| KDM | Endpoint | Model | n | Events | Effect | 95% CI | P | Null |
|---|---|---|---|---|---|---|---|---|
| KDM9 | ADL6 limitation, Wave 7 | Survey-weighted quasibinomial regression | 7,998 | 359 | 1.649 | 1.339, 2.032 | < 0.001 | 1 |
| KDM7 | ADL6 limitation, Wave 7 | Survey-weighted quasibinomial regression | 8,358 | 391 | 1.373 | 1.171, 1.612 | < 0.001 | 1 |
| KDM5 | ADL6 limitation, Wave 7 | Survey-weighted quasibinomial regression | 11,139 | 492 | 1.707 | 1.469, 1.984 | < 0.001 | 1 |
| KDM9 | ADL6 limitation, Wave 8 | Survey-weighted quasibinomial regression | 5,542 | 341 | 1.815 | 1.450, 2.272 | < 0.001 | 1 |
| KDM7 | ADL6 limitation, Wave 8 | Survey-weighted quasibinomial regression | 5,774 | 367 | 1.462 | 1.187, 1.800 | < 0.001 | 1 |
| KDM5 | ADL6 limitation, Wave 8 | Survey-weighted quasibinomial regression | 7,687 | 483 | 1.523 | 1.240, 1.871 | < 0.001 | 1 |
| KDM9 | ADL6 limitation, Wave 9 | Survey-weighted quasibinomial regression | 6,354 | 458 | 1.513 | 1.196, 1.913 | < 0.001 | 1 |
| KDM7 | ADL6 limitation, Wave 9 | Survey-weighted quasibinomial regression | 6,619 | 496 | 1.297 | 1.069, 1.573 | 0.008 | 1 |
| KDM5 | ADL6 limitation, Wave 9 | Survey-weighted quasibinomial regression | 8,767 | 649 | 1.606 | 1.355, 1.903 | < 0.001 | 1 |
| KDM9 | IADL9 limitation, Wave 7 | Survey-weighted quasibinomial regression | 7,664 | 543 | 1.763 | 1.424, 2.184 | < 0.001 | 1 |
| KDM7 | IADL9 limitation, Wave 7 | Survey-weighted quasibinomial regression | 7,984 | 586 | 1.256 | 1.020, 1.548 | 0.032 | 1 |
| KDM5 | IADL9 limitation, Wave 7 | Survey-weighted quasibinomial regression | 10,689 | 709 | 1.604 | 1.367, 1.881 | < 0.001 | 1 |
| KDM9 | IADL9 limitation, Wave 8 | Survey-weighted quasibinomial regression | 5,316 | 513 | 1.476 | 1.131, 1.928 | 0.004 | 1 |
| KDM7 | IADL9 limitation, Wave 8 | Survey-weighted quasibinomial regression | 5,526 | 552 | 1.284 | 1.030, 1.601 | 0.026 | 1 |
| KDM5 | IADL9 limitation, Wave 8 | Survey-weighted quasibinomial regression | 7,389 | 717 | 1.501 | 1.232, 1.828 | < 0.001 | 1 |

Continued on next page

Table S1 continued

| KDM | Endpoint | Model | n | Events | Effect | 95% CI | P | Null |
|---|---|---|---|---|---|---|---|---|
| KDM9 | IADL9 limitation, Wave 9 | Survey-weighted quasibinomial regression | 6,128 | 673 | 1.467 | 1.143, 1.881 | 0.003 | 1 |
| KDM7 | IADL9 limitation, Wave 9 | Survey-weighted quasibinomial regression | 6,365 | 728 | 1.293 | 1.049, 1.595 | 0.016 | 1 |
| KDM5 | IADL9 limitation, Wave 9 | Survey-weighted quasibinomial regression | 8,462 | 967 | 1.521 | 1.238, 1.869 | $< 0.001$ | 1 |
| KDM9 | Multidomain cognition, Wave 9 | Survey-weighted Gaussian regression | 6,676 | – | -0.075 | -0.140, -0.010 | 0.024 | 0 |
| KDM7 | Multidomain cognition, Wave 9 | Survey-weighted Gaussian regression | 6,963 | – | -0.077 | -0.128, -0.027 | 0.003 | 0 |
| KDM5 | Multidomain cognition, Wave 9 | Survey-weighted Gaussian regression | 9,193 | – | -0.057 | -0.103, -0.011 | 0.016 | 0 |
| KDM9 | Mortality through follow-up | Survey-weighted Cox regression | 9,219 | 560 | 1.919 | 1.647, 2.236 | $< 0.001$ | 1 |
| KDM7 | Mortality through follow-up | Survey-weighted Cox regression | 9,711 | 623 | 1.436 | 1.249, 1.651 | $< 0.001$ | 1 |
| KDM5 | Mortality through follow-up | Survey-weighted Cox regression | 12,862 | 811 | 1.741 | 1.503, 2.016 | $< 0.001$ | 1 |

*Note.* Functional endpoints use the six-item ADL and nine-item IADL measures; incident limitation denotes no baseline limitation and any limitation at follow-up. Each KDM measure uses its own outcome-eligible sample. All effect estimates correspond to a 1-SD higher KDM age-acceleration score. Effect is an odds ratio or hazard ratio for ratio-scale models and a regression coefficient for multidomain cognition. Confidence intervals are 95%. P values are unadjusted and should be interpreted relative to the displayed null.

**Table S2:** Pairwise correlations among KDM age-acceleration scores

| Score pair | n | Pearson r (unweighted) | Pearson r (official DBS weighted) |
|---|---|---|---|
| KDM5–KDM7 | 10,397 | 0.693 | 0.692 |
| KDM5–KDM9 | 10,043 | 0.912 | 0.904 |
| KDM7–KDM9 | 10,043 | 0.660 | 0.640 |

*Note.* Pearson correlations use the age-residualized, within-sex standardized KDM acceleration scores, pooled across 12 SHARE countries. Each row uses respondents with both scores available; n is the unweighted number of respondents. Weighted correlations use the official DBS weight (cciw_dbs) on the same pairwise-complete sample, without country normalization or selection reweighting. The correlations are descriptive pairwise measures of overlap among the KDM scores. Restricting all pairs to the common three-score sample (n = 10,043) gives KDM5–KDM7 r = 0.689 unweighted and 0.682 weighted; the other two pairs are unchanged.

**Table S3:** Pooled SES-outcome associations

| KDM | Exposure contrast | Est. | 95% CI | Unadjusted P (coefficient) | BH P (specification) | n | ESS | Countries |
|---|---|---|---|---|---|---|---|---|
| KDM9 | Respondent education: +1 weighted SD | -0.098 | -0.157, -0.039 | 0.001 | 0.001 | 10,043 | 1,186 | 12 |
| KDM7 | Respondent education: +1 weighted SD | -0.107 | -0.159, -0.054 | < 0.001 | < 0.001 | 10,581 | 1,267 | 12 |
| KDM5 | Respondent education: +1 weighted SD | -0.076 | -0.124, -0.029 | 0.002 | 0.002 | 14,015 | 1,867 | 12 |
| KDM9 | Highest-parent education: +1 weighted SD | -0.066 | -0.114, -0.019 | 0.006 | 0.006 | 9,321 | 1,143 | 11 |
| KDM7 | Highest-parent education: +1 weighted SD | -0.075 | -0.133, -0.016 | 0.012 | 0.012 | 9,797 | 1,215 | 11 |
| KDM5 | Highest-parent education: +1 weighted SD | -0.072 | -0.114, -0.030 | < 0.001 | < 0.001 | 12,974 | 1,773 | 11 |
| KDM9 | Wealth rank: +10 percentiles | -0.076 | -0.095, -0.057 | < 0.001 | < 0.001 | 10,003 | 1,173 | 12 |
| KDM7 | Wealth rank: +10 percentiles | -0.061 | -0.080, -0.043 | < 0.001 | < 0.001 | 10,537 | 1,255 | 12 |
| KDM5 | Wealth rank: +10 percentiles | -0.072 | -0.089, -0.055 | < 0.001 | < 0.001 | 13,961 | 1,852 | 12 |
| KDM9 | Income rank: +10 percentiles | -0.074 | -0.092, -0.055 | < 0.001 | < 0.001 | 8,877 | 1,118 | 12 |
| KDM7 | Income rank: +10 percentiles | -0.060 | -0.079, -0.040 | < 0.001 | < 0.001 | 9,354 | 1,192 | 12 |
| KDM5 | Income rank: +10 percentiles | -0.065 | -0.081, -0.048 | < 0.001 | < 0.001 | 12,383 | 1,745 | 12 |
| KDM9 | Residualized educational-rank mobility: +1 weighted SD | -0.119 | -0.162, -0.076 | < 0.001 | < 0.001 | 9,321 | 1,143 | 11 |
| KDM7 | Residualized educational-rank mobility: +1 weighted SD | -0.121 | -0.166, -0.076 | < 0.001 | < 0.001 | 9,797 | 1,215 | 11 |
| KDM5 | Residualized educational-rank mobility: +1 weighted SD | -0.083 | -0.127, -0.040 | < 0.001 | < 0.001 | 12,974 | 1,773 | 11 |
| KDM9 | Parental-origin rank: +10 percentiles | -0.006 | -0.024, 0.012 | 0.509 | < 0.001 | 9,321 | 1,143 | 11 |

Continued on next page

Table S3 continued

| KDM | Exposure contrast | Est. | 95% CI | Unadjusted P (coefficient) | BH P (specification) | n | ESS | Countries |
|---|---|---|---|---|---|---|---|---|
| KDM9 | Respondent-destination rank: +10 percentiles | -0.044 | -0.061, -0.027 | < 0.001 | < 0.001 | 9,321 | 1,143 | 11 |
| KDM7 | Parental-origin rank: +10 percentiles | -0.009 | -0.030, 0.011 | 0.378 | < 0.001 | 9,797 | 1,215 | 11 |
| KDM7 | Respondent-destination rank: +10 percentiles | -0.045 | -0.063, -0.028 | < 0.001 | < 0.001 | 9,797 | 1,215 | 11 |
| KDM5 | Parental-origin rank: +10 percentiles | -0.013 | -0.029, 0.004 | 0.135 | < 0.001 | 12,974 | 1,773 | 11 |
| KDM5 | Respondent-destination rank: +10 percentiles | -0.031 | -0.047, -0.014 | < 0.001 | < 0.001 | 12,974 | 1,773 | 11 |

*Note.* Estimates are differences in KDM age acceleration in SD units for the displayed exposure contrast. Unadjusted P is coefficient-specific. BH-adjusted P values were calculated across the six SES specifications within each outcome; the parental-origin and respondent-destination coefficient rows share one adjusted 2-df block-test P value. ESS denotes weighted effective sample size.

**Table S4:** Magnitude and direction of country-specific SES associations

| KDM | Exposure contrast | Countries | Pooled | Country slope range | Inverse estimates | CI below zero | Tau |
|---|---|---|---|---|---|---|---|
| KDM9 | Respondent education: +1 weighted SD | 12 | -0.098 | -0.213 to 0.054 | 11/12 | 5/12 | 0.072 |
| KDM7 | Respondent education: +1 weighted SD | 12 | -0.107 | -0.198 to -0.005 | 12/12 | 5/12 | 0.033 |
| KDM5 | Respondent education: +1 weighted SD | 12 | -0.076 | -0.184 to 0.003 | 11/12 | 6/12 | 0.050 |
| KDM9 | Highest-parent education: +1 weighted SD | 11 | -0.066 | -0.191 to 0.030 | 9/11 | 6/11 | 0.009 |
| KDM7 | Highest-parent education: +1 weighted SD | 11 | -0.075 | -0.344 to 0.039 | 9/11 | 6/11 | < 0.001 |
| KDM5 | Highest-parent education: +1 weighted SD | 11 | -0.072 | -0.253 to 0.006 | 10/11 | 6/11 | 0.015 |
| KDM9 | Wealth rank: +10 percentiles | 12 | -0.076 | -0.089 to -0.002 | 12/12 | 9/12 | < 0.001 |
| KDM7 | Wealth rank: +10 percentiles | 12 | -0.061 | -0.073 to 0.010 | 11/12 | 9/12 | 0.015 |
| KDM5 | Wealth rank: +10 percentiles | 12 | -0.072 | -0.096 to 0.016 | 11/12 | 10/12 | 0.006 |
| KDM9 | Income rank: +10 percentiles | 12 | -0.074 | -0.115 to 0.030 | 11/12 | 10/12 | 0.022 |
| KDM7 | Income rank: +10 percentiles | 12 | -0.060 | -0.097 to 0.008 | 10/12 | 9/12 | 0.018 |
| KDM5 | Income rank: +10 percentiles | 12 | -0.065 | -0.096 to 0.000 | 11/12 | 10/12 | 0.009 |
| KDM9 | Residualized educational-rank mobility: +1 weighted SD | 11 | -0.119 | -0.202 to 0.020 | 10/11 | 8/11 | 0.052 |
| KDM7 | Residualized educational-rank mobility: +1 weighted SD | 11 | -0.121 | -0.235 to 0.024 | 9/11 | 4/11 | 0.037 |
| KDM5 | Residualized educational-rank mobility: +1 weighted SD | 11 | -0.083 | -0.172 to 0.037 | 10/11 | 6/11 | 0.040 |
| KDM9 | Parental-origin rank: +10 percentiles | 11 | -0.006 | -0.061 to 0.051 | 9/11 | 1/11 | < 0.001 |
| KDM9 | Respondent-destination rank: +10 percentiles | 11 | -0.044 | -0.080 to 0.007 | 9/11 | 8/11 | 0.020 |

Continued on next page

Table S4 continued

| **KDM** | **Exposure contrast** | **Countries** | **Pooled** | **Country slope range** | **Inverse estimates** | **CI below zero** | **Tau** |
|---|---|---|---|---|---|---|---|
| KDM7 | Parental-origin rank: +10 percentiles | 11 | -0.009 | -0.145 to 0.029 | 8/11 | 4/11 | $< 0.001$ |
| KDM7 | Respondent-destination rank: +10 percentiles | 11 | -0.045 | -0.093 to 0.038 | 9/11 | 4/11 | 0.015 |
| KDM5 | Parental-origin rank: +10 percentiles | 11 | -0.013 | -0.100 to 0.027 | 8/11 | 3/11 | $< 0.001$ |
| KDM5 | Respondent-destination rank: +10 percentiles | 11 | -0.031 | -0.066 to 0.018 | 10/11 | 6/11 | 0.015 |

*Note.* Pooled coefficients, country slopes, and tau use KDM acceleration SD units per displayed SES increment. Country slopes are linear combinations of the pooled country-varying model. Range is the minimum and maximum country point estimate; it includes sampling variation. The "Inverse estimates" column counts negative point estimates; the "CI below zero" column counts country-specific pointwise 95% intervals wholly below zero, without multiplicity adjustment. Tau is the square root of the moderator-free full-covariance REML variance; parental-origin and respondent-destination contrasts have separate variance components. Tau is an estimate with uncertainty, especially with 11 or 12 countries. Values below 0.0005 are displayed as <0.001 and do not establish exact homogeneity. The parental-origin and respondent-destination rows correspond to two contrasts within one omnibus specification. Education and mobility increments use country-specific weighted SDs; economic, parental-origin, and respondent-destination ranks use 10-percentile increments. Ranges across different exposure increments are not directly comparable.

**Table S5:** Sex and baseline-age heterogeneity

| KDM | SES specification | Interaction | F | df | Unadjusted P | BH(6) | BH(12) |
|---|---|---|---|---|---|---|---|
| KDM9 | Respondent education | Sex | 0.11 | 1, 6620.0 | 0.737 | 0.737 | 0.778 |
| KDM9 | Highest-parent education | Sex | 0.32 | 1, 6175.0 | 0.572 | 0.737 | 0.763 |
| KDM9 | Wealth rank | Sex | 0.17 | 1, 296.4 | 0.681 | 0.737 | 0.778 |
| KDM9 | Income rank | Sex | 1.32 | 1, 1725.2 | 0.251 | 0.501 | 0.667 |
| KDM9 | Residualized educational-rank mobility | Sex | 1.73 | 1, 6175.0 | 0.189 | 0.501 | 0.667 |
| KDM9 | Joint origin–destination | Sex | 2.21 | 2, 6163.0 | 0.110 | 0.501 | 0.667 |
| KDM7 | Respondent education | Sex | 4.51 | 1, 6938.0 | 0.034 | 0.101 | 0.202 |
| KDM7 | Highest-parent education | Sex | 4.65 | 1, 6454.0 | 0.031 | 0.101 | 0.202 |
| KDM7 | Wealth rank | Sex | 3.04 | 1, 2314.0 | 0.081 | 0.142 | 0.284 |
| KDM7 | Income rank | Sex | 0.25 | 1, 2759.9 | 0.615 | 0.663 | 0.869 |
| KDM7 | Residualized educational-rank mobility | Sex | 0.19 | 1, 6454.0 | 0.663 | 0.663 | 0.869 |
| KDM7 | Joint origin–destination | Sex | 2.36 | 2, 6442.0 | 0.095 | 0.142 | 0.284 |
| KDM5 | Respondent education | Sex | 2.76 | 1, 8593.0 | 0.096 | 0.579 | 0.758 |
| KDM5 | Highest-parent education | Sex | 1.22 | 1, 7969.0 | 0.270 | 0.695 | 0.758 |
| KDM5 | Wealth rank | Sex | 0.04 | 1, 3250.6 | 0.845 | 0.845 | 0.845 |
| KDM5 | Income rank | Sex | 0.72 | 1, 446.9 | 0.395 | 0.695 | 0.758 |
| KDM5 | Residualized educational-rank mobility | Sex | 0.18 | 1, 7969.0 | 0.670 | 0.804 | 0.804 |
| KDM5 | Joint origin–destination | Sex | 0.77 | 2, 7957.0 | 0.463 | 0.695 | 0.758 |
| KDM9 | Respondent education | Baseline age, linear | 0.48 | 1, 6620.0 | 0.489 | 0.587 | 0.734 |
| KDM9 | Highest-parent education | Baseline age, linear | 1.82 | 1, 6175.0 | 0.177 | 0.583 | 0.667 |
| KDM9 | Wealth rank | Baseline age, linear | 1.02 | 1, 3545.3 | 0.312 | 0.583 | 0.667 |
| KDM9 | Income rank | Baseline age, linear | 0.75 | 1, 95.9 | 0.389 | 0.583 | 0.667 |

Continued on next page

Table S5 continued

| KDM | SES specification | Interaction | F | df | Unadjusted P | BH(6) | BH(12) |
|---|---|---|---|---|---|---|---|
| KDM9 | Residualized educational-rank mobility | Baseline age, linear | 0.08 | 1, 6175.0 | 0.778 | 0.778 | 0.778 |
| KDM9 | Joint origin–destination | Baseline age, linear | 1.00 | 2, 6163.0 | 0.367 | 0.583 | 0.667 |
| KDM7 | Respondent education | Baseline age, linear | 0.00 | 1, 6938.0 | 0.971 | 0.971 | 0.971 |
| KDM7 | Highest-parent education | Baseline age, linear | 1.68 | 1, 6454.0 | 0.195 | 0.846 | 0.467 |
| KDM7 | Wealth rank | Baseline age, linear | 0.01 | 1, 1939.9 | 0.906 | 0.971 | 0.971 |
| KDM7 | Income rank | Baseline age, linear | 0.12 | 1, 372.4 | 0.724 | 0.971 | 0.869 |
| KDM7 | Residualized educational-rank mobility | Baseline age, linear | 0.64 | 1, 6454.0 | 0.423 | 0.846 | 0.726 |
| KDM7 | Joint origin–destination | Baseline age, linear | 0.98 | 2, 6442.0 | 0.374 | 0.846 | 0.726 |
| KDM5 | Respondent education | Baseline age, linear | 1.60 | 1, 8593.0 | 0.205 | 0.742 | 0.758 |
| KDM5 | Highest-parent education | Baseline age, linear | 0.73 | 1, 7969.0 | 0.393 | 0.742 | 0.758 |
| KDM5 | Wealth rank | Baseline age, linear | 0.11 | 1, 4967.1 | 0.742 | 0.742 | 0.809 |
| KDM5 | Income rank | Baseline age, linear | 0.61 | 1, 480.2 | 0.434 | 0.742 | 0.758 |
| KDM5 | Residualized educational-rank mobility | Baseline age, linear | 0.44 | 1, 7969.0 | 0.505 | 0.742 | 0.758 |
| KDM5 | Joint origin–destination | Baseline age, linear | 0.45 | 2, 7957.0 | 0.641 | 0.742 | 0.804 |
| KDM9 | Respondent education | Baseline age, spline | 0.65 | 3, 6618.0 | 0.581 | 0.581 | – |
| KDM9 | Highest-parent education | Baseline age, spline | 1.04 | 3, 6173.0 | 0.373 | 0.519 | – |

Continued on next page

Table S5 continued

| KDM | SES specification | Interaction | F | df | Unadjusted P | BH(6) | BH(12) |
|---|---|---|---|---|---|---|---|
| KDM9 | Wealth rank | Baseline age, spline | 0.92 | 3, 1390.6 | 0.432 | 0.519 | – |
| KDM9 | Income rank | Baseline age, spline | 1.51 | 3, 280.7 | 0.213 | 0.519 | – |
| KDM9 | Residualized educational-rank mobility | Baseline age, spline | 1.51 | 3, 6173.0 | 0.211 | 0.519 | – |
| KDM9 | Joint origin–destination | Baseline age, spline | 1.16 | 6, 6159.0 | 0.325 | 0.519 | – |
| KDM7 | Respondent education | Baseline age, spline | 0.17 | 3, 6936.0 | 0.918 | 0.926 | – |
| KDM7 | Highest-parent education | Baseline age, spline | 1.21 | 3, 6452.0 | 0.305 | 0.766 | – |
| KDM7 | Wealth rank | Baseline age, spline | 0.16 | 3, 3231.2 | 0.926 | 0.926 | – |
| KDM7 | Income rank | Baseline age, spline | 0.77 | 3, 1382.0 | 0.511 | 0.766 | – |
| KDM7 | Residualized educational-rank mobility | Baseline age, spline | 0.86 | 3, 6452.0 | 0.463 | 0.766 | – |
| KDM7 | Joint origin–destination | Baseline age, spline | 0.97 | 6, 6438.0 | 0.444 | 0.766 | – |

Continued on next page

Table S5 continued

| KDM | SES specification | Interaction | F | df | Unadjusted P | BH(6) | BH(12) |
|---|---|---|---|---|---|---|---|
| KDM5 | Respondent education | Baseline age, spline | 0.66 | 3, 8591.0 | 0.575 | 0.989 | – |
| KDM5 | Highest-parent education | Baseline age, spline | 0.48 | 3, 7967.0 | 0.693 | 0.989 | – |
| KDM5 | Wealth rank | Baseline age, spline | 0.04 | 3, 1668.2 | 0.989 | 0.989 | – |
| KDM5 | Income rank | Baseline age, spline | 0.43 | 3, 638.4 | 0.728 | 0.989 | – |
| KDM5 | Residualized educational-rank mobility | Baseline age, spline | 0.31 | 3, 7967.0 | 0.821 | 0.989 | – |
| KDM5 | Joint origin–destination | Baseline age, spline | 0.41 | 6, 7953.0 | 0.872 | 0.989 | – |

*Note.* BH(6) denotes Benjamini–Hochberg adjustment across six SES tests within each outcome and interaction type. BH(12) denotes adjustment across the 12 tests formed by the six sex and six linear baseline-age tests within each outcome. Because all baseline measurements were obtained in the same survey wave, age contrasts cannot be separated from birth-cohort differences and may also be affected by selective survival.

**Table S6:** KDM9 sex-specific and baseline-age margins

| Panel | Exposure contrast | Sex | Baseline age | Estimate (KDM SD) | 95% CI | P |
|---|---|---|---|---|---|---|
| A | Respondent education: +1 weighted SD | Female | 62.5 | -0.105 | -0.179, -0.032 | 0.005 |
| A | Respondent education: +1 weighted SD | Male | 62.5 | -0.087 | -0.171, -0.003 | 0.044 |
| A | Highest-parent education: +1 weighted SD | Female | 62.5 | -0.080 | -0.151, -0.008 | 0.029 |
| A | Highest-parent education: +1 weighted SD | Male | 62.5 | -0.050 | -0.117, 0.016 | 0.138 |
| A | Wealth rank: +10 percentiles | Female | 62.5 | -0.080 | -0.104, -0.055 | $< 0.001$ |
| A | Wealth rank: +10 percentiles | Male | 62.5 | -0.072 | -0.101, -0.042 | $< 0.001$ |
| A | Income rank: +10 percentiles | Female | 62.5 | -0.064 | -0.090, -0.038 | $< 0.001$ |
| A | Income rank: +10 percentiles | Male | 62.5 | -0.085 | -0.110, -0.060 | $< 0.001$ |
| A | Residualized educational-rank mobility: +1 weighted SD | Female | 62.5 | -0.092 | -0.149, -0.034 | 0.002 |
| A | Residualized educational-rank mobility: +1 weighted SD | Male | 62.5 | -0.148 | -0.211, -0.086 | $< 0.001$ |
| A | Parental-origin rank: +10 percentiles | Female | 62.5 | -0.019 | -0.043, 0.005 | 0.121 |
| A | Parental-origin rank: +10 percentiles | Male | 62.5 | 0.011 | -0.016, 0.037 | 0.422 |
| A | Respondent-destination rank: +10 percentiles | Female | 62.5 | -0.032 | -0.055, -0.010 | 0.005 |
| A | Respondent-destination rank: +10 percentiles | Male | 62.5 | -0.058 | -0.082, -0.035 | $< 0.001$ |

Continued on next page

Table S6 continued

| Panel | Exposure contrast | Sex | Base-line age | Estimate (KDM SD) | 95% CI | P |
|---|---|---|---|---|---|---|
| B | Respondent education: +1 weighted SD | Male | 50.0 | 0.006 | -0.189, 0.201 | 0.952 |
| B | Respondent education: +1 weighted SD | Male | 55.0 | -0.063 | -0.164, 0.038 | 0.223 |
| B | Respondent education: +1 weighted SD | Male | 60.0 | -0.114 | -0.215, -0.014 | 0.026 |
| B | Respondent education: +1 weighted SD | Male | 65.0 | -0.133 | -0.208, -0.059 | < 0.001 |
| B | Respondent education: +1 weighted SD | Male | 70.0 | -0.118 | -0.204, -0.032 | 0.007 |
| B | Respondent education: +1 weighted SD | Male | 75.0 | -0.081 | -0.220, 0.059 | 0.257 |
| B | Highest-parent education: +1 weighted SD | Male | 50.0 | -0.123 | -0.322, 0.075 | 0.224 |
| B | Highest-parent education: +1 weighted SD | Male | 55.0 | -0.038 | -0.115, 0.039 | 0.335 |
| B | Highest-parent education: +1 weighted SD | Male | 60.0 | -0.016 | -0.106, 0.073 | 0.720 |
| B | Highest-parent education: +1 weighted SD | Male | 65.0 | -0.086 | -0.149, -0.022 | 0.008 |
| B | Highest-parent education: +1 weighted SD | Male | 70.0 | -0.130 | -0.210, -0.050 | 0.001 |
| B | Highest-parent education: +1 weighted SD | Male | 75.0 | -0.116 | -0.240, 0.008 | 0.066 |
| B | Wealth rank: +10 percentiles | Male | 50.0 | -0.017 | -0.095, 0.060 | 0.657 |
| B | Wealth rank: +10 percentiles | Male | 55.0 | -0.066 | -0.098, -0.034 | < 0.001 |
| B | Wealth rank: +10 percentiles | Male | 60.0 | -0.088 | -0.120, -0.057 | < 0.001 |
| B | Wealth rank: +10 percentiles | Male | 65.0 | -0.075 | -0.099, -0.051 | < 0.001 |
| B | Wealth rank: +10 percentiles | Male | 70.0 | -0.077 | -0.105, -0.049 | < 0.001 |
| B | Wealth rank: +10 percentiles | Male | 75.0 | -0.109 | -0.185, -0.032 | 0.005 |
| B | Income rank: +10 percentiles | Male | 50.0 | -0.007 | -0.093, 0.079 | 0.870 |
| B | Income rank: +10 percentiles | Male | 55.0 | -0.061 | -0.090, -0.031 | < 0.001 |
| B | Income rank: +10 percentiles | Male | 60.0 | -0.092 | -0.124, -0.061 | < 0.001 |
| B | Income rank: +10 percentiles | Male | 65.0 | -0.089 | -0.112, -0.066 | < 0.001 |
| B | Income rank: +10 percentiles | Male | 70.0 | -0.075 | -0.108, -0.041 | < 0.001 |
| B | Income rank: +10 percentiles | Male | 75.0 | -0.066 | -0.119, -0.012 | 0.016 |
| B | Residualized educational-rank mobility: +1 weighted SD | Male | 50.0 | 0.047 | -0.129, 0.223 | 0.600 |
| B | Residualized educational-rank mobility: +1 weighted SD | Male | 55.0 | -0.122 | -0.193, -0.051 | < 0.001 |
| B | Residualized educational-rank mobility: +1 weighted SD | Male | 60.0 | -0.187 | -0.265, -0.109 | < 0.001 |
| B | Residualized educational-rank mobility: +1 weighted SD | Male | 65.0 | -0.105 | -0.184, -0.026 | 0.009 |

Continued on next page

Table S6 continued

| Panel | Exposure contrast | Sex | Base-line age | Estimate (KDM SD) | 95% CI | P |
|---|---|---|---|---|---|---|
| B | Residualized educational-rank mobility: +1 weighted SD | Male | 70.0 | -0.069 | -0.153, 0.016 | 0.110 |
| B | Residualized educational-rank mobility: +1 weighted SD | Male | 75.0 | -0.131 | -0.313, 0.050 | 0.155 |
| B | Parental-origin rank: +10 percentiles | Male | 50.0 | -0.040 | -0.112, 0.032 | 0.277 |
| B | Parental-origin rank: +10 percentiles | Male | 55.0 | 0.005 | -0.022, 0.033 | 0.706 |
| B | Parental-origin rank: +10 percentiles | Male | 60.0 | 0.019 | -0.013, 0.051 | 0.241 |
| B | Parental-origin rank: +10 percentiles | Male | 65.0 | -0.012 | -0.038, 0.013 | 0.338 |
| B | Parental-origin rank: +10 percentiles | Male | 70.0 | -0.033 | -0.063, -0.002 | 0.036 |
| B | Parental-origin rank: +10 percentiles | Male | 75.0 | -0.025 | -0.081, 0.031 | 0.375 |
| B | Respondent-destination rank: +10 percentiles | Male | 50.0 | 0.012 | -0.054, 0.077 | 0.725 |
| B | Respondent-destination rank: +10 percentiles | Male | 55.0 | -0.046 | -0.073, -0.019 | 0.001 |
| B | Respondent-destination rank: +10 percentiles | Male | 60.0 | -0.068 | -0.098, -0.039 | $< 0.001$ |
| B | Respondent-destination rank: +10 percentiles | Male | 65.0 | -0.041 | -0.070, -0.011 | 0.007 |
| B | Respondent-destination rank: +10 percentiles | Male | 70.0 | -0.029 | -0.060, 0.002 | 0.063 |
| B | Respondent-destination rank: +10 percentiles | Male | 75.0 | -0.051 | -0.116, 0.014 | 0.124 |

*Note.* Estimates are KDM9 age-acceleration SD differences for the displayed exposure contrast. Panel A reports sex-specific margins at age 62.5 years; Panel B reports male baseline-age spline margins at ages 50–75 years. Because all baseline measurements were obtained in the same survey wave, age contrasts cannot be separated from birth-cohort differences and may also be affected by selective survival. Global interaction tests are reported in Table S5.

**Table S7:** Country-context moderation

| KDM | SES specification | Moderator | CR2 F | df | CR2 P | CWB P | BH CR2 | BH CWB | Stability note |
|---|---|---|---|---|---|---|---|---|---|
| KDM9 | Respondent education | Social spending | 0.00 | 1, 3.6 | 0.959 | 0.952 | 0.959 | 0.952 | Limited effective df |
| KDM9 | Respondent education | Disposable-income Gini | 0.82 | 1, 6.7 | 0.396 | 0.401 | 0.808 | 0.818 | No diagnostic flag |
| KDM9 | Respondent education | Low-earner pension | 0.20 | 1, 3.2 | 0.683 | 0.615 | 0.808 | 0.818 | Limited effective df |
| KDM9 | Respondent education | Out-of-pocket health share | 2.06 | 1, 3.4 | 0.236 | 0.421 | 0.808 | 0.818 | Limited effective df |
| KDM9 | Respondent education | GDP per capita | 0.15 | 1, 4.1 | 0.715 | 0.728 | 0.808 | 0.818 | No diagnostic flag |
| KDM9 | Respondent education | Unemployment | 0.51 | 1, 1.9 | 0.551 | 0.712 | 0.808 | 0.818 | Limited effective df |
| KDM9 | Highest-parent education | Social spending | 1.18 | 1, 2.8 | 0.363 | 0.242 | 0.808 | 0.818 | Limited effective df |
| KDM9 | Highest-parent education | Disposable-income Gini | 0.05 | 1, 4.3 | 0.840 | 0.825 | 0.890 | 0.874 | Country leverage concern |
| KDM9 | Highest-parent education | Low-earner pension | 0.24 | 1, 2.3 | 0.668 | 0.613 | 0.808 | 0.818 | Limited effective df |
| KDM9 | Highest-parent education | Out-of-pocket health share | 0.28 | 1, 4.8 | 0.619 | 0.600 | 0.808 | 0.818 | No diagnostic flag |
| KDM9 | Highest-parent education | GDP per capita | 1.52 | 1, 2.4 | 0.324 | 0.190 | 0.808 | 0.818 | Limited effective df |
| KDM9 | Highest-parent education | Unemployment | 0.02 | 1, 1.9 | 0.906 | 0.921 | 0.932 | 0.947 | Limited effective df |
| KDM9 | Wealth rank | Social spending | 4.25 | 1, 2.6 | 0.147 | 0.022 | 0.808 | 0.792 | Limited effective df |
| KDM9 | Wealth rank | Disposable-income Gini | 1.15 | 1, 4.8 | 0.336 | 0.311 | 0.808 | 0.818 | No diagnostic flag |
| KDM9 | Wealth rank | Low-earner pension | 0.18 | 1, 2.4 | 0.704 | 0.728 | 0.808 | 0.818 | Limited effective df |
| KDM9 | Wealth rank | Out-of-pocket health share | 2.09 | 1, 5.0 | 0.207 | 0.256 | 0.808 | 0.818 | No diagnostic flag |
| KDM9 | Wealth rank | GDP per capita | 1.42 | 1, 3.1 | 0.316 | 0.276 | 0.808 | 0.818 | Limited effective df |
| KDM9 | Wealth rank | Unemployment | 0.53 | 1, 2.2 | 0.536 | 0.819 | 0.808 | 0.874 | Limited effective df |
| KDM9 | Income rank | Social spending | 1.26 | 1, 4.0 | 0.324 | 0.265 | 0.808 | 0.818 | No diagnostic flag |
| KDM9 | Income rank | Disposable-income Gini | 0.49 | 1, 6.7 | 0.508 | 0.563 | 0.808 | 0.818 | No diagnostic flag |
| KDM9 | Income rank | Low-earner pension | 0.15 | 1, 3.4 | 0.718 | 0.679 | 0.808 | 0.818 | Limited effective df |

Continued on next page

Table S7 continued

| KDM | SES specification | Moderator | CR2 F | df | CR2 P | CWB P | BH CR2 | BH CWB | Stability note |
|---|---|---|---|---|---|---|---|---|---|
| KDM9 | Income rank | Out-of-pocket health share | 2.01 | 1, 3.8 | 0.233 | 0.412 | 0.808 | 0.818 | Limited effective df |
| KDM9 | Income rank | GDP per capita | 0.56 | 1, 4.3 | 0.494 | 0.720 | 0.808 | 0.818 | No diagnostic flag |
| KDM9 | Income rank | Unemployment | 0.40 | 1, 2.0 | 0.590 | 0.681 | 0.808 | 0.818 | Limited effective df |
| KDM9 | Residualized educational-rank mobility | Social spending | 0.57 | 1, 3.3 | 0.501 | 0.648 | 0.808 | 0.818 | Limited effective df |
| KDM9 | Residualized educational-rank mobility | Disposable-income Gini | 0.29 | 1, 5.3 | 0.612 | 0.599 | 0.808 | 0.818 | No diagnostic flag |
| KDM9 | Residualized educational-rank mobility | Low-earner pension | 0.83 | 1, 2.7 | 0.437 | 0.316 | 0.808 | 0.818 | Limited effective df |
| KDM9 | Residualized educational-rank mobility | Out-of-pocket health share | 1.42 | 1, 6.5 | 0.275 | 0.278 | 0.808 | 0.818 | No diagnostic flag |
| KDM9 | Residualized educational-rank mobility | GDP per capita | 4.23 | 1, 2.8 | 0.137 | 0.174 | 0.808 | 0.818 | Limited effective df |
| KDM9 | Residualized educational-rank mobility | Unemployment | 1.43 | 1, 1.9 | 0.358 | 0.344 | 0.808 | 0.818 | Limited effective df |
| KDM9 | Joint origin–destination | Social spending | 1.14 | 2, 2.1 | 0.462 | 0.562 | 0.808 | 0.818 | Limited effective df |
| KDM9 | Joint origin–destination | Disposable-income Gini | 1.68 | 2, 3.7 | 0.304 | 0.216 | 0.808 | 0.818 | Limited effective df |
| KDM9 | Joint origin–destination | Low-earner pension | 0.34 | 2, 1.4 | 0.756 | 0.499 | 0.825 | 0.818 | Limited effective df |
| KDM9 | Joint origin–destination | Out-of-pocket health share | 1.24 | 2, 4.5 | 0.372 | 0.329 | 0.808 | 0.818 | Country leverage concern |
| KDM9 | Joint origin–destination | GDP per capita | 2.61 | 2, 1.5 | 0.320 | 0.231 | 0.808 | 0.818 | Limited effective df |
| KDM9 | Joint origin–destination | Unemployment | 0.49 | 2, 1.1 | 0.705 | 0.686 | 0.808 | 0.818 | Limited effective df |
| KDM7 | Respondent education | Social spending | 3.95 | 1, 3.1 | 0.138 | 0.090 | 0.611 | 0.465 | Limited effective df |

Continued on next page

Table S7 continued

| KDM | SES specification | Moderator | CR2 F | df | CR2 P | CWB P | BH CR2 | BH CWB | Stability note |
|---|---|---|---|---|---|---|---|---|---|
| KDM7 | Respondent education | Disposable-income Gini | 0.12 | 1, 6.3 | 0.745 | 0.738 | 1.000 | 1.000 | Leave-one-country-out unstable |
| KDM7 | Respondent education | Low-earner pension | 0.91 | 1, 2.8 | 0.416 | 0.380 | 0.859 | 0.719 | Limited effective df |
| KDM7 | Respondent education | Out-of-pocket health share | 3.13 | 1, 4.1 | 0.150 | 0.175 | 0.611 | 0.544 | No diagnostic flag |
| KDM7 | Respondent education | GDP per capita | 0.00 | 1, 4.1 | 0.983 | 0.985 | 1.000 | 1.000 | Leave-one-country-out unstable |
| KDM7 | Respondent education | Unemployment | 0.00 | 1, 1.8 | 0.978 | 0.971 | 1.000 | 1.000 | Limited effective df |
| KDM7 | Highest-parent education | Social spending | 0.00 | 1, 2.9 | 1.000 | 1.000 | 1.000 | 1.000 | Limited effective df |
| KDM7 | Highest-parent education | Disposable-income Gini | 0.24 | 1, 4.6 | 0.648 | 0.708 | 1.000 | 1.000 | No diagnostic flag |
| KDM7 | Highest-parent education | Low-earner pension | 1.38 | 1, 2.3 | 0.346 | 0.160 | 0.830 | 0.544 | Limited effective df |
| KDM7 | Highest-parent education | Out-of-pocket health share | 0.03 | 1, 4.6 | 0.880 | 0.889 | 1.000 | 1.000 | Country leverage concern |
| KDM7 | Highest-parent education | GDP per capita | 3.17 | 1, 2.6 | 0.187 | 0.081 | 0.611 | 0.465 | Limited effective df |
| KDM7 | Highest-parent education | Unemployment | 0.00 | 1, 1.9 | 0.996 | 0.996 | 1.000 | 1.000 | Limited effective df |
| KDM7 | Wealth rank | Social spending | 3.18 | 1, 3.5 | 0.160 | 0.036 | 0.611 | 0.461 | Limited effective df |
| KDM7 | Wealth rank | Disposable-income Gini | 0.39 | 1, 6.2 | 0.555 | 0.557 | 0.999 | 0.956 | No diagnostic flag |
| KDM7 | Wealth rank | Low-earner pension | 0.83 | 1, 3.0 | 0.430 | 0.225 | 0.859 | 0.544 | Limited effective df |
| KDM7 | Wealth rank | Out-of-pocket health share | 2.07 | 1, 4.3 | 0.219 | 0.376 | 0.658 | 0.719 | No diagnostic flag |
| KDM7 | Wealth rank | GDP per capita | 6.53 | 1, 3.8 | 0.066 | 0.197 | 0.611 | 0.544 | Limited effective df |
| KDM7 | Wealth rank | Unemployment | 0.22 | 1, 1.9 | 0.687 | 0.822 | 1.000 | 1.000 | Limited effective df |
| KDM7 | Income rank | Social spending | 1.76 | 1, 3.7 | 0.260 | 0.150 | 0.720 | 0.544 | Limited effective df |
| KDM7 | Income rank | Disposable-income Gini | 0.32 | 1, 6.2 | 0.594 | 0.600 | 1.000 | 0.981 | No diagnostic flag |
| KDM7 | Income rank | Low-earner pension | 0.00 | 1, 3.2 | 0.999 | 1.000 | 1.000 | 1.000 | Limited effective df |

Continued on next page

Table S7 continued

| KDM | SES specification | Moderator | CR2 F | df | CR2 P | CWB P | BH CR2 | BH CWB | Stability note |
|---|---|---|---|---|---|---|---|---|---|
| KDM7 | Income rank | Out-of-pocket health share | 0.57 | 1, 4.1 | 0.493 | 0.521 | 0.934 | 0.939 | No diagnostic flag |
| KDM7 | Income rank | GDP per capita | 3.34 | 1, 4.2 | 0.138 | 0.211 | 0.611 | 0.544 | No diagnostic flag |
| KDM7 | Income rank | Unemployment | 0.00 | 1, 2.0 | 0.978 | 0.910 | 1.000 | 1.000 | Limited effective df |
| KDM7 | Residualized educational-rank mobility | Social spending | 16.67 | 1, 2.8 | 0.029 | 0.025 | 0.611 | 0.461 | Limited effective df |
| KDM7 | Residualized educational-rank mobility | Disposable-income Gini | 2.44 | 1, 5.2 | 0.177 | 0.172 | 0.611 | 0.544 | No diagnostic flag |
| KDM7 | Residualized educational-rank mobility | Low-earner pension | 0.01 | 1, 2.6 | 0.929 | 0.924 | 1.000 | 1.000 | Limited effective df |
| KDM7 | Residualized educational-rank mobility | Out-of-pocket health share | 7.04 | 1, 5.9 | 0.039 | 0.064 | 0.611 | 0.461 | Inference-method sensitive |
| KDM7 | Residualized educational-rank mobility | GDP per capita | 0.00 | 1, 3.2 | 0.976 | 0.980 | 1.000 | 1.000 | Limited effective df |
| KDM7 | Residualized educational-rank mobility | Unemployment | 1.06 | 1, 1.8 | 0.419 | 0.347 | 0.859 | 0.719 | Limited effective df |
| KDM7 | Joint origin–destination | Social spending | 6.82 | 2, 2.3 | 0.106 | 0.042 | 0.611 | 0.461 | Limited effective df |
| KDM7 | Joint origin–destination | Disposable-income Gini | 1.67 | 2, 4.2 | 0.292 | 0.227 | 0.750 | 0.544 | Country leverage concern |
| KDM7 | Joint origin–destination | Low-earner pension | 0.65 | 2, 1.5 | 0.625 | 0.370 | 1.000 | 0.719 | Limited effective df |
| KDM7 | Joint origin–destination | Out-of-pocket health share | 3.58 | 2, 5.0 | 0.108 | 0.060 | 0.611 | 0.461 | Country leverage concern |
| KDM7 | Joint origin–destination | GDP per capita | 0.20 | 2, 2.0 | 0.830 | 0.823 | 1.000 | 1.000 | Limited effective df |
| KDM7 | Joint origin–destination | Unemployment | 0.26 | 2, 0.9 | 0.813 | 0.761 | 1.000 | 1.000 | Limited effective df |
| KDM5 | Respondent education | Social spending | 0.25 | 1, 3.8 | 0.646 | 0.610 | 0.874 | 0.839 | Limited effective df |

Continued on next page

Table S7 continued

| KDM | SES specification | Moderator | CR2 F | df | CR2 P | CWB P | BH CR2 | BH CWB | Stability note |
|---|---|---|---|---|---|---|---|---|---|
| KDM5 | Respondent education | Disposable-income Gini | 0.41 | 1, 6.7 | 0.542 | 0.528 | 0.874 | 0.839 | No diagnostic flag |
| KDM5 | Respondent education | Low-earner pension | 0.56 | 1, 3.3 | 0.505 | 0.419 | 0.874 | 0.794 | Limited effective df |
| KDM5 | Respondent education | Out-of-pocket health share | 1.60 | 1, 3.6 | 0.281 | 0.370 | 0.848 | 0.794 | Limited effective df |
| KDM5 | Respondent education | GDP per capita | 0.89 | 1, 4.2 | 0.398 | 0.403 | 0.874 | 0.794 | No diagnostic flag |
| KDM5 | Respondent education | Unemployment | 0.33 | 1, 2.0 | 0.623 | 0.597 | 0.874 | 0.839 | Limited effective df |
| KDM5 | Highest-parent education | Social spending | 2.99 | 1, 3.3 | 0.175 | 0.035 | 0.848 | 0.639 | Limited effective df |
| KDM5 | Highest-parent education | Disposable-income Gini | 1.42 | 1, 4.9 | 0.288 | 0.262 | 0.848 | 0.794 | No diagnostic flag |
| KDM5 | Highest-parent education | Low-earner pension | 0.08 | 1, 2.5 | 0.798 | 0.774 | 0.898 | 0.871 | Limited effective df |
| KDM5 | Highest-parent education | Out-of-pocket health share | 0.36 | 1, 5.8 | 0.572 | 0.556 | 0.874 | 0.839 | No diagnostic flag |
| KDM5 | Highest-parent education | GDP per capita | 0.56 | 1, 3.0 | 0.508 | 0.367 | 0.874 | 0.794 | Limited effective df |
| KDM5 | Highest-parent education | Unemployment | 0.01 | 1, 2.1 | 0.928 | 0.981 | 0.982 | 0.999 | Limited effective df |
| KDM5 | Wealth rank | Social spending | 1.31 | 1, 2.9 | 0.336 | 0.269 | 0.848 | 0.794 | Limited effective df |
| KDM5 | Wealth rank | Disposable-income Gini | 1.87 | 1, 5.6 | 0.224 | 0.201 | 0.848 | 0.794 | No diagnostic flag |
| KDM5 | Wealth rank | Low-earner pension | 0.02 | 1, 2.7 | 0.906 | 0.878 | 0.982 | 0.958 | Limited effective df |
| KDM5 | Wealth rank | Out-of-pocket health share | 8.30 | 1, 5.1 | 0.034 | 0.080 | 0.848 | 0.794 | Inference-method sensitive |
| KDM5 | Wealth rank | GDP per capita | 5.26 | 1, 3.5 | 0.094 | 0.015 | 0.848 | 0.522 | Limited effective df |
| KDM5 | Wealth rank | Unemployment | 0.46 | 1, 2.2 | 0.561 | 0.641 | 0.874 | 0.839 | Limited effective df |
| KDM5 | Income rank | Social spending | 1.14 | 1, 3.5 | 0.353 | 0.370 | 0.848 | 0.794 | Limited effective df |
| KDM5 | Income rank | Disposable-income Gini | 0.21 | 1, 6.0 | 0.661 | 0.650 | 0.874 | 0.839 | No diagnostic flag |
| KDM5 | Income rank | Low-earner pension | 0.00 | 1, 3.0 | 0.985 | 0.979 | 0.999 | 0.999 | Limited effective df |
| KDM5 | Income rank | Out-of-pocket health share | 4.84 | 1, 4.5 | 0.085 | 0.160 | 0.848 | 0.794 | No diagnostic flag |

Continued on next page

Table S7 continued

| KDM | SES specification | Moderator | CR2 F | df | CR2 P | CWB P | BH CR2 | BH CWB | Stability note |
|---|---|---|---|---|---|---|---|---|---|
| KDM5 | Income rank | GDP per capita | 0.20 | 1, 4.1 | 0.680 | 0.675 | 0.874 | 0.839 | No diagnostic flag |
| KDM5 | Income rank | Unemployment | 0.29 | 1, 2.1 | 0.639 | 0.626 | 0.874 | 0.839 | Limited effective df |
| KDM5 | Residualized educational-rank mobility | Social spending | 0.11 | 1, 3.7 | 0.756 | 0.730 | 0.898 | 0.871 | Limited effective df |
| KDM5 | Residualized educational-rank mobility | Disposable-income Gini | 1.13 | 1, 5.4 | 0.333 | 0.305 | 0.848 | 0.794 | No diagnostic flag |
| KDM5 | Residualized educational-rank mobility | Low-earner pension | 0.25 | 1, 2.7 | 0.658 | 0.625 | 0.874 | 0.839 | Limited effective df |
| KDM5 | Residualized educational-rank mobility | Out-of-pocket health share | 1.76 | 1, 6.5 | 0.229 | 0.214 | 0.848 | 0.794 | No diagnostic flag |
| KDM5 | Residualized educational-rank mobility | GDP per capita | 5.00 | 1, 3.0 | 0.111 | 0.167 | 0.848 | 0.794 | Limited effective df |
| KDM5 | Residualized educational-rank mobility | Unemployment | 0.00 | 1, 2.2 | 0.999 | 0.999 | 0.999 | 0.999 | Limited effective df |
| KDM5 | Joint origin–destination | Social spending | 1.68 | 2, 2.5 | 0.345 | 0.265 | 0.848 | 0.794 | Limited effective df |
| KDM5 | Joint origin–destination | Disposable-income Gini | 1.58 | 2, 4.0 | 0.312 | 0.291 | 0.848 | 0.794 | Country leverage concern |
| KDM5 | Joint origin–destination | Low-earner pension | 0.26 | 2, 1.5 | 0.797 | 0.669 | 0.898 | 0.839 | Limited effective df |
| KDM5 | Joint origin–destination | Out-of-pocket health share | 1.01 | 2, 5.2 | 0.426 | 0.412 | 0.874 | 0.794 | Country leverage concern |
| KDM5 | Joint origin–destination | GDP per capita | 3.12 | 2, 1.9 | 0.248 | 0.127 | 0.848 | 0.794 | Limited effective df |
| KDM5 | Joint origin–destination | Unemployment | 0.34 | 2, 1.4 | 0.758 | 0.766 | 0.898 | 0.871 | Limited effective df |

*Note.* CR2 uses small-sample robust meta-regression inference; CWB is the country wild-bootstrap P value. BH columns report P values adjusted across the 36 moderator tests within each outcome. The stability note summarizes effective degrees of freedom, country leverage, leave-one-country-out direction, and inference-method sensitivity; the underlying continuous statistics are reported in the table where applicable. The models describe associations between country context and country-specific SES gradients.

**Table S8:** Paired CWB covariance sensitivity

| KDM | SES specification | Moderator | Full F | Full CR2 P | Full CWB P | Full BH | Block F | Block CR2 P | Block CWB P | Block BH |
|---|---|---|---|---|---|---|---|---|---|---|
| KDM9 | Respondent education | Social spending | 0.00 | 0.959 | 0.952 | 0.952 | 0.00 | 0.957 | 0.954 | 0.954 |
| KDM9 | Respondent education | Disposable-income Gini | 0.82 | 0.396 | 0.401 | 0.818 | 0.82 | 0.397 | 0.399 | 0.816 |
| KDM9 | Respondent education | Low-earner pension | 0.20 | 0.683 | 0.615 | 0.818 | 0.20 | 0.685 | 0.620 | 0.816 |
| KDM9 | Respondent education | Out-of-pocket health share | 2.06 | 0.236 | 0.421 | 0.818 | 2.06 | 0.236 | 0.421 | 0.816 |
| KDM9 | Respondent education | GDP per capita | 0.15 | 0.715 | 0.728 | 0.818 | 0.16 | 0.713 | 0.725 | 0.816 |
| KDM9 | Respondent education | Unemployment | 0.51 | 0.551 | 0.712 | 0.818 | 0.51 | 0.551 | 0.715 | 0.816 |
| KDM9 | Highest-parent education | Social spending | 1.18 | 0.363 | 0.242 | 0.818 | 1.06 | 0.383 | 0.244 | 0.816 |
| KDM9 | Highest-parent education | Disposable-income Gini | 0.05 | 0.840 | 0.825 | 0.874 | 0.05 | 0.837 | 0.818 | 0.866 |
| KDM9 | Highest-parent education | Low-earner pension | 0.24 | 0.668 | 0.613 | 0.818 | 0.21 | 0.686 | 0.622 | 0.816 |
| KDM9 | Highest-parent education | Out-of-pocket health share | 0.28 | 0.619 | 0.600 | 0.818 | 0.24 | 0.643 | 0.613 | 0.816 |
| KDM9 | Highest-parent education | GDP per capita | 1.52 | 0.324 | 0.190 | 0.818 | 1.48 | 0.332 | 0.180 | 0.816 |
| KDM9 | Highest-parent education | Unemployment | 0.02 | 0.906 | 0.921 | 0.947 | 0.02 | 0.910 | 0.921 | 0.948 |
| KDM9 | Wealth rank | Social spending | 4.25 | 0.147 | 0.022 | 0.792 | 4.17 | 0.141 | 0.022 | 0.810 |
| KDM9 | Wealth rank | Disposable-income Gini | 1.15 | 0.336 | 0.311 | 0.818 | 1.19 | 0.327 | 0.300 | 0.816 |
| KDM9 | Wealth rank | Low-earner pension | 0.18 | 0.704 | 0.728 | 0.818 | 0.21 | 0.688 | 0.624 | 0.816 |
| KDM9 | Wealth rank | Out-of-pocket health share | 2.09 | 0.207 | 0.256 | 0.818 | 2.17 | 0.198 | 0.239 | 0.816 |
| KDM9 | Wealth rank | GDP per capita | 1.42 | 0.316 | 0.276 | 0.818 | 1.36 | 0.324 | 0.276 | 0.816 |
| KDM9 | Wealth rank | Unemployment | 0.53 | 0.536 | 0.819 | 0.874 | 0.58 | 0.517 | 0.777 | 0.848 |
| KDM9 | Income rank | Social spending | 1.26 | 0.324 | 0.265 | 0.818 | 1.25 | 0.327 | 0.257 | 0.816 |
| KDM9 | Income rank | Disposable-income Gini | 0.49 | 0.508 | 0.563 | 0.818 | 0.48 | 0.510 | 0.575 | 0.816 |
| KDM9 | Income rank | Low-earner pension | 0.15 | 0.718 | 0.679 | 0.818 | 0.17 | 0.704 | 0.669 | 0.816 |
| KDM9 | Income rank | Out-of-pocket health share | 2.01 | 0.233 | 0.412 | 0.818 | 1.90 | 0.245 | 0.461 | 0.816 |
| KDM9 | Income rank | GDP per capita | 0.56 | 0.494 | 0.720 | 0.818 | 0.57 | 0.488 | 0.720 | 0.816 |
| KDM9 | Income rank | Unemployment | 0.40 | 0.590 | 0.681 | 0.818 | 0.43 | 0.580 | 0.676 | 0.816 |
| KDM9 | Residualized educational-rank mobility | Social spending | 0.57 | 0.501 | 0.648 | 0.818 | 0.57 | 0.501 | 0.649 | 0.816 |
| KDM9 | Residualized educational-rank mobility | Disposable-income Gini | 0.29 | 0.612 | 0.599 | 0.818 | 0.29 | 0.611 | 0.599 | 0.816 |

Continued on next page

Table S8 continued

| KDM | SES specification | Moderator | Full F | Full CR2 P | Full CWB P | Full BH | Block F | Block CR2 P | Block CWB P | Block BH |
|---|---|---|---|---|---|---|---|---|---|---|
| KDM9 | Residualized educational-rank mobility | Low-earner pension | 0.83 | 0.437 | 0.316 | 0.818 | 0.81 | 0.442 | 0.322 | 0.816 |
| KDM9 | Residualized educational-rank mobility | Out-of-pocket health share | 1.42 | 0.275 | 0.278 | 0.818 | 1.44 | 0.273 | 0.272 | 0.816 |
| KDM9 | Residualized educational-rank mobility | GDP per capita | 4.23 | 0.137 | 0.174 | 0.818 | 4.32 | 0.134 | 0.173 | 0.816 |
| KDM9 | Residualized educational-rank mobility | Unemployment | 1.43 | 0.358 | 0.344 | 0.818 | 1.43 | 0.357 | 0.344 | 0.816 |
| KDM9 | Joint origin–destination | Social spending | 1.14 | 0.462 | 0.562 | 0.818 | 1.11 | 0.468 | 0.562 | 0.816 |
| KDM9 | Joint origin–destination | Disposable-income Gini | 1.68 | 0.304 | 0.216 | 0.818 | 1.65 | 0.309 | 0.218 | 0.816 |
| KDM9 | Joint origin–destination | Low-earner pension | 0.34 | 0.756 | 0.499 | 0.818 | 0.33 | 0.761 | 0.520 | 0.816 |
| KDM9 | Joint origin–destination | Out-of-pocket health share | 1.24 | 0.372 | 0.329 | 0.818 | 1.22 | 0.377 | 0.330 | 0.816 |
| KDM9 | Joint origin–destination | GDP per capita | 2.61 | 0.320 | 0.231 | 0.818 | 2.79 | 0.308 | 0.198 | 0.816 |
| KDM9 | Joint origin–destination | Unemployment | 0.49 | 0.705 | 0.686 | 0.818 | 0.51 | 0.697 | 0.682 | 0.816 |

Continued on next page

Table S8 continued

| KDM | SES specification | Moderator | Full F | Full CR2 P | Full CWB P | Full BH | Block F | Block CR2 P | Block CWB P | Block BH |
|---|---|---|---|---|---|---|---|---|---|---|
| KDM7 | Respondent education | Social spending | 3.95 | 0.138 | 0.090 | 0.465 | 4.06 | 0.138 | 0.090 | 0.463 |
| KDM7 | Respondent education | Disposable-income Gini | 0.12 | 0.745 | 0.738 | 1.000 | 0.10 | 0.758 | 0.745 | 1.000 |
| KDM7 | Respondent education | Low-earner pension | 0.91 | 0.416 | 0.380 | 0.719 | 0.91 | 0.415 | 0.371 | 0.703 |
| KDM7 | Respondent education | Out-of-pocket health share | 3.13 | 0.150 | 0.175 | 0.544 | 3.14 | 0.149 | 0.175 | 0.546 |
| KDM7 | Respondent education | GDP per capita | 0.00 | 0.983 | 0.985 | 1.000 | 0.00 | 0.989 | 0.986 | 1.000 |
| KDM7 | Respondent education | Unemployment | 0.00 | 0.978 | 0.971 | 1.000 | 0.00 | 0.978 | 0.968 | 1.000 |
| KDM7 | Highest-parent education | Social spending | 0.00 | 1.000 | 1.000 | 1.000 | 0.00 | 0.978 | 0.979 | 1.000 |
| KDM7 | Highest-parent education | Disposable-income Gini | 0.24 | 0.648 | 0.708 | 1.000 | 0.28 | 0.620 | 0.658 | 1.000 |
| KDM7 | Highest-parent education | Low-earner pension | 1.38 | 0.346 | 0.160 | 0.544 | 1.23 | 0.369 | 0.215 | 0.546 |
| KDM7 | Highest-parent education | Out-of-pocket health share | 0.03 | 0.880 | 0.889 | 1.000 | 0.02 | 0.890 | 0.906 | 1.000 |
| KDM7 | Highest-parent education | GDP per capita | 3.17 | 0.187 | 0.081 | 0.465 | 3.37 | 0.178 | 0.061 | 0.381 |
| KDM7 | Highest-parent education | Unemployment | 0.00 | 0.996 | 0.996 | 1.000 | 0.00 | 0.992 | 0.988 | 1.000 |
| KDM7 | Wealth rank | Social spending | 3.18 | 0.160 | 0.036 | 0.461 | 3.11 | 0.163 | 0.041 | 0.381 |
| KDM7 | Wealth rank | Disposable-income Gini | 0.39 | 0.555 | 0.557 | 0.956 | 0.44 | 0.533 | 0.555 | 0.952 |
| KDM7 | Wealth rank | Low-earner pension | 0.83 | 0.430 | 0.225 | 0.544 | 0.85 | 0.425 | 0.228 | 0.546 |
| KDM7 | Wealth rank | Out-of-pocket health share | 2.07 | 0.219 | 0.376 | 0.719 | 2.13 | 0.212 | 0.359 | 0.703 |
| KDM7 | Wealth rank | GDP per capita | 6.53 | 0.066 | 0.197 | 0.544 | 6.29 | 0.069 | 0.182 | 0.546 |
| KDM7 | Wealth rank | Unemployment | 0.22 | 0.687 | 0.822 | 1.000 | 0.23 | 0.679 | 0.798 | 1.000 |
| KDM7 | Income rank | Social spending | 1.76 | 0.260 | 0.150 | 0.544 | 1.75 | 0.261 | 0.153 | 0.546 |
| KDM7 | Income rank | Disposable-income Gini | 0.32 | 0.594 | 0.600 | 0.981 | 0.31 | 0.599 | 0.602 | 0.984 |
| KDM7 | Income rank | Low-earner pension | 0.00 | 0.999 | 1.000 | 1.000 | 0.00 | 1.000 | 1.000 | 1.000 |
| KDM7 | Income rank | Out-of-pocket health share | 0.57 | 0.493 | 0.521 | 0.939 | 0.59 | 0.484 | 0.516 | 0.930 |
| KDM7 | Income rank | GDP per capita | 3.34 | 0.138 | 0.211 | 0.544 | 3.57 | 0.128 | 0.203 | 0.546 |
| KDM7 | Income rank | Unemployment | 0.00 | 0.978 | 0.910 | 1.000 | 0.00 | 0.987 | 0.948 | 1.000 |
| KDM7 | Residualized educational-rank mobility | Social spending | 16.67 | 0.029 | 0.025 | 0.461 | 16.42 | 0.031 | 0.025 | 0.381 |
| KDM7 | Residualized educational-rank mobility | Disposable-income Gini | 2.44 | 0.177 | 0.172 | 0.544 | 2.45 | 0.177 | 0.172 | 0.546 |

Continued on next page

Table S8 continued

| KDM | SES specification | Moderator | Full F | Full CR2 P | Full CWB P | Full BH | Block F | Block CR2 P | Block CWB P | Block BH |
|---|---|---|---|---|---|---|---|---|---|---|
| KDM7 | Residualized educational-rank mobility | Low-earner pension | 0.01 | 0.929 | 0.924 | 1.000 | 0.01 | 0.923 | 0.922 | 1.000 |
| KDM7 | Residualized educational-rank mobility | Out-of-pocket health share | 7.04 | 0.039 | 0.064 | 0.461 | 6.78 | 0.042 | 0.064 | 0.381 |
| KDM7 | Residualized educational-rank mobility | GDP per capita | 0.00 | 0.976 | 0.980 | 1.000 | 0.00 | 0.990 | 0.994 | 1.000 |
| KDM7 | Residualized educational-rank mobility | Unemployment | 1.06 | 0.419 | 0.347 | 0.719 | 1.06 | 0.419 | 0.346 | 0.703 |
| KDM7 | Joint origin–destination | Social spending | 6.82 | 0.106 | 0.042 | 0.461 | 6.73 | 0.108 | 0.042 | 0.381 |
| KDM7 | Joint origin–destination | Disposable-income Gini | 1.67 | 0.292 | 0.227 | 0.544 | 1.66 | 0.295 | 0.226 | 0.546 |
| KDM7 | Joint origin–destination | Low-earner pension | 0.65 | 0.625 | 0.370 | 0.719 | 0.65 | 0.625 | 0.368 | 0.703 |
| KDM7 | Joint origin–destination | Out-of-pocket health share | 3.58 | 0.108 | 0.060 | 0.461 | 3.46 | 0.114 | 0.061 | 0.381 |
| KDM7 | Joint origin–destination | GDP per capita | 0.20 | 0.830 | 0.823 | 1.000 | 0.21 | 0.829 | 0.829 | 1.000 |
| KDM7 | Joint origin–destination | Unemployment | 0.26 | 0.813 | 0.761 | 1.000 | 0.26 | 0.814 | 0.765 | 1.000 |

Continued on next page

Table S8 continued

| KDM | SES specification | Moderator | Full F | Full CR2 P | Full CWB P | Full BH | Block F | Block CR2 P | Block CWB P | Block BH |
|---|---|---|---|---|---|---|---|---|---|---|
| KDM5 | Respondent education | Social spending | 0.25 | 0.646 | 0.610 | 0.839 | 0.24 | 0.649 | 0.610 | 0.845 |
| KDM5 | Respondent education | Disposable-income Gini | 0.41 | 0.542 | 0.528 | 0.839 | 0.41 | 0.542 | 0.531 | 0.845 |
| KDM5 | Respondent education | Low-earner pension | 0.56 | 0.505 | 0.419 | 0.794 | 0.54 | 0.510 | 0.419 | 0.795 |
| KDM5 | Respondent education | Out-of-pocket health share | 1.60 | 0.281 | 0.370 | 0.794 | 1.61 | 0.280 | 0.373 | 0.795 |
| KDM5 | Respondent education | GDP per capita | 0.89 | 0.398 | 0.403 | 0.794 | 0.89 | 0.396 | 0.401 | 0.795 |
| KDM5 | Respondent education | Unemployment | 0.33 | 0.623 | 0.597 | 0.839 | 0.33 | 0.622 | 0.602 | 0.845 |
| KDM5 | Highest-parent education | Social spending | 2.99 | 0.175 | 0.035 | 0.639 | 2.91 | 0.179 | 0.035 | 0.639 |
| KDM5 | Highest-parent education | Disposable-income Gini | 1.42 | 0.288 | 0.262 | 0.794 | 1.39 | 0.293 | 0.274 | 0.795 |
| KDM5 | Highest-parent education | Low-earner pension | 0.08 | 0.798 | 0.774 | 0.871 | 0.07 | 0.814 | 0.792 | 0.891 |
| KDM5 | Highest-parent education | Out-of-pocket health share | 0.36 | 0.572 | 0.556 | 0.839 | 0.33 | 0.587 | 0.580 | 0.845 |
| KDM5 | Highest-parent education | GDP per capita | 0.56 | 0.508 | 0.367 | 0.794 | 0.53 | 0.520 | 0.373 | 0.795 |
| KDM5 | Highest-parent education | Unemployment | 0.01 | 0.928 | 0.981 | 0.999 | 0.01 | 0.940 | 0.984 | 0.990 |
| KDM5 | Wealth rank | Social spending | 1.31 | 0.336 | 0.269 | 0.794 | 1.28 | 0.337 | 0.237 | 0.795 |
| KDM5 | Wealth rank | Disposable-income Gini | 1.87 | 0.224 | 0.201 | 0.794 | 2.05 | 0.205 | 0.206 | 0.795 |
| KDM5 | Wealth rank | Low-earner pension | 0.02 | 0.906 | 0.878 | 0.958 | 0.02 | 0.896 | 0.869 | 0.947 |
| KDM5 | Wealth rank | Out-of-pocket health share | 8.30 | 0.034 | 0.080 | 0.794 | 8.19 | 0.033 | 0.070 | 0.795 |
| KDM5 | Wealth rank | GDP per capita | 5.26 | 0.094 | 0.015 | 0.522 | 4.72 | 0.105 | 0.016 | 0.576 |
| KDM5 | Wealth rank | Unemployment | 0.46 | 0.561 | 0.641 | 0.839 | 0.59 | 0.513 | 0.589 | 0.845 |
| KDM5 | Income rank | Social spending | 1.14 | 0.353 | 0.370 | 0.794 | 1.03 | 0.379 | 0.394 | 0.795 |
| KDM5 | Income rank | Disposable-income Gini | 0.21 | 0.661 | 0.650 | 0.839 | 0.20 | 0.670 | 0.665 | 0.845 |
| KDM5 | Income rank | Low-earner pension | 0.00 | 0.985 | 0.979 | 0.999 | 0.00 | 0.985 | 0.981 | 0.990 |
| KDM5 | Income rank | Out-of-pocket health share | 4.84 | 0.085 | 0.160 | 0.794 | 4.39 | 0.096 | 0.180 | 0.795 |
| KDM5 | Income rank | GDP per capita | 0.20 | 0.680 | 0.675 | 0.839 | 0.20 | 0.675 | 0.681 | 0.845 |
| KDM5 | Income rank | Unemployment | 0.29 | 0.639 | 0.626 | 0.839 | 0.32 | 0.626 | 0.623 | 0.845 |
| KDM5 | Residualized educational-rank mobility | Social spending | 0.11 | 0.756 | 0.730 | 0.871 | 0.11 | 0.753 | 0.730 | 0.875 |
| KDM5 | Residualized educational-rank mobility | Disposable-income Gini | 1.13 | 0.333 | 0.305 | 0.794 | 1.13 | 0.334 | 0.305 | 0.795 |

Continued on next page

Table S8 continued

| KDM | SES specification | Moderator | Full F | Full CR2 P | Full CWB P | Full BH | Block F | Block CR2 P | Block CWB P | Block BH |
|---|---|---|---|---|---|---|---|---|---|---|
| KDM5 | Residualized educational-rank mobility | Low-earner pension | 0.25 | 0.658 | 0.625 | 0.839 | 0.24 | 0.659 | 0.625 | 0.845 |
| KDM5 | Residualized educational-rank mobility | Out-of-pocket health share | 1.76 | 0.229 | 0.214 | 0.794 | 1.76 | 0.229 | 0.214 | 0.795 |
| KDM5 | Residualized educational-rank mobility | GDP per capita | 5.00 | 0.111 | 0.167 | 0.794 | 4.97 | 0.111 | 0.167 | 0.795 |
| KDM5 | Residualized educational-rank mobility | Unemployment | 0.00 | 0.999 | 0.999 | 0.999 | 0.00 | 0.995 | 0.990 | 0.990 |
| KDM5 | Joint origin–destination | Social spending | 1.68 | 0.345 | 0.265 | 0.794 | 1.66 | 0.347 | 0.269 | 0.795 |
| KDM5 | Joint origin–destination | Disposable-income Gini | 1.58 | 0.312 | 0.291 | 0.794 | 1.56 | 0.314 | 0.291 | 0.795 |
| KDM5 | Joint origin–destination | Low-earner pension | 0.26 | 0.797 | 0.669 | 0.839 | 0.26 | 0.800 | 0.673 | 0.845 |
| KDM5 | Joint origin–destination | Out-of-pocket health share | 1.01 | 0.426 | 0.412 | 0.794 | 1.00 | 0.427 | 0.413 | 0.795 |
| KDM5 | Joint origin–destination | GDP per capita | 3.12 | 0.248 | 0.127 | 0.794 | 3.14 | 0.247 | 0.126 | 0.795 |
| KDM5 | Joint origin–destination | Unemployment | 0.34 | 0.758 | 0.766 | 0.871 | 0.35 | 0.756 | 0.767 | 0.891 |

*Note.* Full retains the complete country-slope sampling covariance, including off-country elements; Block retains all variances and within-country covariance but sets off-country elements to zero. Across 108 paired cells, maximum absolute differences in $P$ values were 0.047 for CR2 and 0.104 for CWB. No raw $P < 0.05$ classification changed for either method, and no CWB BH-adjusted decision changed. BH columns report CWB adjustment within each KDM outcome (36 tests). This same-engine paired sensitivity is not an independent validation of Full.

**Table S9:** Data support for descriptive country association profiles

| Country | Exact components | SES-outcome cells |
|---|---|---|
| Belgium | 21/21 | 18/18 |
| Denmark | 21/21 | 18/18 |
| Estonia | 21/21 | 18/18 |
| France | 21/21 | 18/18 |
| Germany | 21/21 | 18/18 |
| Greece | 9/21 | 9/18 |
| Israel | 21/21 | 18/18 |
| Italy | 21/21 | 18/18 |
| Slovenia | 21/21 | 18/18 |
| Spain | 21/21 | 18/18 |
| Sweden | 21/21 | 18/18 |
| Switzerland | 21/21 | 18/18 |

*Note.* A complete country profile required all 21 exact coefficients and all 18 SES–outcome cells. Eleven countries met this requirement; Greece lacked the parental-education-based components and was not included in the 11-country complete-profile display.

**Table S10:** Country association profiles across socioeconomic dimensions

| **Country** | **Respondent education** | **Highest-parent education** | **Wealth rank** | **Income rank** | **Residualized educational-rank mobility** | **Joint origin–destination** |
|---|---|---|---|---|---|---|
| Belgium | 0.52 | 0.20 | 0.42 | 0.03 | 0.26 | 0.12 |
| Denmark | 0.40 | -0.13 | 0.09 | -0.05 | 0.43 | 0.14 |
| Estonia | 0.56 | -0.09 | -0.60 | -0.19 | 0.26 | 0.16 |
| France | -0.03 | -0.74 | 0.46 | 1.00 | 1.57 | 0.11 |
| Germany | 0.02 | 0.12 | 0.74 | 0.57 | 0.31 | 0.15 |
| Israel | 0.58 | 1.88 | -2.32 | -1.37 | -1.40 | 0.20 |
| Italy | -1.30 | -1.25 | 0.38 | -0.39 | -0.93 | -0.84 |
| Slovenia | 0.64 | -0.34 | -0.42 | -0.84 | 0.76 | 0.06 |
| Spain | 1.02 | 1.23 | 1.11 | 1.80 | 0.53 | 0.76 |
| Sweden | -1.08 | -0.76 | 0.14 | -0.01 | -0.56 | -0.39 |
| Switzerland | -1.34 | -0.12 | -0.01 | -0.54 | -1.23 | -0.47 |

*Note.* Each SES column is an equal-weight mean of country-standardized coefficients across the three KDM outcomes. Coefficients were multiplied by $-1$ before standardization, so larger values indicate a stronger inverse association between socioeconomic advantage and KDM acceleration. Origin and destination values were averaged within KDM outcome before aggregation. Countries are shown alphabetically; the profile values are descriptive and do not incorporate coefficient precision.

**Table S11:** Selection-weight sensitivity

| KDM | Exposure contrast | Weighting scheme | Est. | 95% CI | P | n | ESS | Countries |
|---|---|---|---|---|---|---|---|---|
| KDM9 | Respondent education: +1 weighted SD | Official DBS | -0.098 | -0.157, -0.039 | 0.001 | 10,043 | 1,186 | 12 |
| KDM9 | Respondent education: +1 weighted SD | Trimmed official DBS | -0.098 | -0.157, -0.039 | 0.001 | 10,043 | 1,193 | 12 |
| KDM9 | Respondent education: +1 weighted SD | SIPW | -0.113 | -0.173, -0.053 | < 0.001 | 10,043 | 948 | 12 |
| KDM9 | Respondent education: +1 weighted SD | Trimmed SIPW | -0.115 | -0.175, -0.055 | < 0.001 | 10,043 | 958 | 12 |
| KDM9 | Respondent education: +1 weighted SD | Unit weights | -0.137 | -0.157, -0.116 | < 0.001 | 10,043 | 10,043 | 12 |
| KDM9 | Highest-parent education: +1 weighted SD | Official DBS | -0.066 | -0.114, -0.019 | 0.006 | 9,321 | 1,143 | 11 |
| KDM9 | Highest-parent education: +1 weighted SD | Trimmed official DBS | -0.066 | -0.114, -0.019 | 0.006 | 9,321 | 1,151 | 11 |
| KDM9 | Highest-parent education: +1 weighted SD | SIPW | -0.075 | -0.124, -0.026 | 0.003 | 9,321 | 885 | 11 |
| KDM9 | Highest-parent education: +1 weighted SD | Trimmed SIPW | -0.072 | -0.120, -0.024 | 0.003 | 9,321 | 895 | 11 |
| KDM9 | Highest-parent education: +1 weighted SD | Unit weights | -0.086 | -0.106, -0.065 | < 0.001 | 9,321 | 9,321 | 11 |
| KDM9 | Wealth rank: +10 percentiles | Official DBS | -0.076 | -0.095, -0.057 | < 0.001 | 10,003 | 1,173 | 12 |
| KDM9 | Wealth rank: +10 percentiles | Trimmed official DBS | -0.076 | -0.095, -0.057 | < 0.001 | 10,003 | 1,180 | 12 |
| KDM9 | Wealth rank: +10 percentiles | SIPW | -0.079 | -0.102, -0.056 | < 0.001 | 10,003 | 940 | 12 |
| KDM9 | Wealth rank: +10 percentiles | Trimmed SIPW | -0.078 | -0.101, -0.055 | < 0.001 | 10,003 | 950 | 12 |
| KDM9 | Wealth rank: +10 percentiles | Unit weights | -0.072 | -0.080, -0.064 | < 0.001 | 10,003 | 10,003 | 12 |
| KDM9 | Income rank: +10 percentiles | Official DBS | -0.074 | -0.092, -0.055 | < 0.001 | 8,877 | 1,118 | 12 |
| KDM9 | Income rank: +10 percentiles | Trimmed official DBS | -0.074 | -0.093, -0.055 | < 0.001 | 8,877 | 1,126 | 12 |
| KDM9 | Income rank: +10 percentiles | SIPW | -0.075 | -0.098, -0.052 | < 0.001 | 8,877 | 866 | 12 |
| KDM9 | Income rank: +10 percentiles | Trimmed SIPW | -0.074 | -0.097, -0.052 | < 0.001 | 8,877 | 874 | 12 |
| KDM9 | Income rank: +10 percentiles | Unit weights | -0.063 | -0.071, -0.055 | < 0.001 | 8,877 | 8,877 | 12 |
| KDM9 | Residualized educational-rank mobility: +1 weighted SD | Official DBS | -0.119 | -0.162, -0.076 | < 0.001 | 9,321 | 1,143 | 11 |
| KDM9 | Residualized educational-rank mobility: +1 weighted SD | Trimmed official DBS | -0.120 | -0.163, -0.077 | < 0.001 | 9,321 | 1,151 | 11 |

Continued on next page

Table S11 continued

| KDM | Exposure contrast | Weighting scheme | Est. | 95% CI | P | n | ESS | Countries |
|---|---|---|---|---|---|---|---|---|
| KDM9 | Residualized educational-rank mobility: +1 weighted SD | SIPW | -0.125 | -0.173, -0.078 | < 0.001 | 9,321 | 885 | 11 |
| KDM9 | Residualized educational-rank mobility: +1 weighted SD | Trimmed SIPW | -0.128 | -0.174, -0.081 | < 0.001 | 9,321 | 895 | 11 |
| KDM9 | Residualized educational-rank mobility: +1 weighted SD | Unit weights | -0.123 | -0.143, -0.103 | < 0.001 | 9,321 | 9,321 | 11 |
| KDM9 | Parental-origin rank: +10 percentiles | Official DBS | -0.006 | -0.024, 0.012 | 0.509 | 9,321 | 1,143 | 11 |
| KDM9 | Parental-origin rank: +10 percentiles | Trimmed official DBS | -0.006 | -0.024, 0.012 | 0.510 | 9,321 | 1,151 | 11 |
| KDM9 | Parental-origin rank: +10 percentiles | SIPW | -0.008 | -0.028, 0.012 | 0.431 | 9,321 | 885 | 11 |
| KDM9 | Parental-origin rank: +10 percentiles | Trimmed SIPW | -0.007 | -0.026, 0.013 | 0.500 | 9,321 | 895 | 11 |
| KDM9 | Parental-origin rank: +10 percentiles | Unit weights | -0.014 | -0.022, -0.006 | < 0.001 | 9,321 | 9,321 | 11 |
| KDM9 | Respondent-destination rank: +10 percentiles | Official DBS | -0.044 | -0.061, -0.027 | < 0.001 | 9,321 | 1,143 | 11 |
| KDM9 | Respondent-destination rank: +10 percentiles | Trimmed official DBS | -0.044 | -0.061, -0.028 | < 0.001 | 9,321 | 1,151 | 11 |
| KDM9 | Respondent-destination rank: +10 percentiles | SIPW | -0.046 | -0.064, -0.028 | < 0.001 | 9,321 | 885 | 11 |
| KDM9 | Respondent-destination rank: +10 percentiles | Trimmed SIPW | -0.047 | -0.065, -0.029 | < 0.001 | 9,321 | 895 | 11 |
| KDM9 | Respondent-destination rank: +10 percentiles | Unit weights | -0.046 | -0.053, -0.038 | < 0.001 | 9,321 | 9,321 | 11 |

Continued on next page

Table S11 continued

| KDM | Exposure contrast | Weighting scheme | Est. | 95% CI | P | n | ESS | Countries |
|---|---|---|---|---|---|---|---|---|
| KDM7 | Respondent education: +1 weighted SD | Official DBS | -0.107 | -0.159, -0.054 | < 0.001 | 10,581 | 1,267 | 12 |
| KDM7 | Respondent education: +1 weighted SD | Trimmed official DBS | -0.107 | -0.160, -0.055 | < 0.001 | 10,581 | 1,275 | 12 |
| KDM7 | Respondent education: +1 weighted SD | SIPW | -0.101 | -0.161, -0.041 | < 0.001 | 10,448 | 928 | 11 |
| KDM7 | Respondent education: +1 weighted SD | Trimmed SIPW | -0.102 | -0.162, -0.043 | < 0.001 | 10,448 | 931 | 11 |
| KDM7 | Respondent education: +1 weighted SD | Unit weights | -0.098 | -0.117, -0.078 | < 0.001 | 10,581 | 10,581 | 12 |
| KDM7 | Highest-parent education: +1 weighted SD | Official DBS | -0.075 | -0.133, -0.016 | 0.012 | 9,797 | 1,215 | 11 |
| KDM7 | Highest-parent education: +1 weighted SD | Trimmed official DBS | -0.075 | -0.134, -0.017 | 0.012 | 9,797 | 1,224 | 11 |
| KDM7 | Highest-parent education: +1 weighted SD | SIPW | -0.078 | -0.142, -0.014 | 0.016 | 9,689 | 867 | 10 |
| KDM7 | Highest-parent education: +1 weighted SD | Trimmed SIPW | -0.078 | -0.142, -0.013 | 0.018 | 9,689 | 870 | 10 |
| KDM7 | Highest-parent education: +1 weighted SD | Unit weights | -0.068 | -0.088, -0.048 | < 0.001 | 9,797 | 9,797 | 11 |
| KDM7 | Wealth rank: +10 percentiles | Official DBS | -0.061 | -0.080, -0.043 | < 0.001 | 10,537 | 1,255 | 12 |
| KDM7 | Wealth rank: +10 percentiles | Trimmed official DBS | -0.062 | -0.079, -0.044 | < 0.001 | 10,537 | 1,262 | 12 |
| KDM7 | Wealth rank: +10 percentiles | SIPW | -0.068 | -0.090, -0.047 | < 0.001 | 10,406 | 920 | 11 |
| KDM7 | Wealth rank: +10 percentiles | Trimmed SIPW | -0.068 | -0.090, -0.047 | < 0.001 | 10,406 | 923 | 11 |
| KDM7 | Wealth rank: +10 percentiles | Unit weights | -0.048 | -0.055, -0.041 | < 0.001 | 10,537 | 10,537 | 12 |
| KDM7 | Income rank: +10 percentiles | Official DBS | -0.060 | -0.079, -0.040 | < 0.001 | 9,354 | 1,192 | 12 |
| KDM7 | Income rank: +10 percentiles | Trimmed official DBS | -0.060 | -0.079, -0.041 | < 0.001 | 9,354 | 1,201 | 12 |
| KDM7 | Income rank: +10 percentiles | SIPW | -0.067 | -0.089, -0.044 | < 0.001 | 9,250 | 853 | 11 |
| KDM7 | Income rank: +10 percentiles | Trimmed SIPW | -0.066 | -0.089, -0.044 | < 0.001 | 9,250 | 856 | 11 |
| KDM7 | Income rank: +10 percentiles | Unit weights | -0.038 | -0.046, -0.031 | < 0.001 | 9,354 | 9,354 | 12 |
| KDM7 | Residualized educational-rank mobility: +1 weighted SD | Official DBS | -0.121 | -0.166, -0.076 | < 0.001 | 9,797 | 1,215 | 11 |
| KDM7 | Residualized educational-rank mobility: +1 weighted SD | Trimmed official DBS | -0.122 | -0.167, -0.076 | < 0.001 | 9,797 | 1,224 | 11 |
| KDM7 | Residualized educational-rank mobility: +1 weighted SD | SIPW | -0.118 | -0.170, -0.067 | < 0.001 | 9,689 | 867 | 10 |

Continued on next page

Table S11 continued

| KDM | Exposure contrast | Weighting scheme | Est. | 95% CI | P | n | ESS | Countries |
|---|---|---|---|---|---|---|---|---|
| KDM7 | Residualized educational-rank mobility: +1 weighted SD | Trimmed SIPW | -0.120 | -0.171, -0.069 | < 0.001 | 9,689 | 870 | 10 |
| KDM7 | Residualized educational-rank mobility: +1 weighted SD | Unit weights | -0.088 | -0.107, -0.069 | < 0.001 | 9,797 | 9,797 | 11 |
| KDM7 | Parental-origin rank: +10 percentiles | Official DBS | -0.009 | -0.030, 0.011 | 0.378 | 9,797 | 1,215 | 11 |
| KDM7 | Parental-origin rank: +10 percentiles | Trimmed official DBS | -0.010 | -0.030, 0.011 | 0.357 | 9,797 | 1,224 | 11 |
| KDM7 | Parental-origin rank: +10 percentiles | SIPW | -0.013 | -0.036, 0.010 | 0.260 | 9,689 | 867 | 10 |
| KDM7 | Parental-origin rank: +10 percentiles | Trimmed SIPW | -0.013 | -0.035, 0.010 | 0.286 | 9,689 | 870 | 10 |
| KDM7 | Parental-origin rank: +10 percentiles | Unit weights | -0.013 | -0.021, -0.006 | < 0.001 | 9,797 | 9,797 | 11 |
| KDM7 | Respondent-destination rank: +10 percentiles | Official DBS | -0.045 | -0.063, -0.028 | < 0.001 | 9,797 | 1,215 | 11 |
| KDM7 | Respondent-destination rank: +10 percentiles | Trimmed official DBS | -0.045 | -0.063, -0.028 | < 0.001 | 9,797 | 1,224 | 11 |
| KDM7 | Respondent-destination rank: +10 percentiles | SIPW | -0.044 | -0.064, -0.024 | < 0.001 | 9,689 | 867 | 10 |
| KDM7 | Respondent-destination rank: +10 percentiles | Trimmed SIPW | -0.045 | -0.064, -0.025 | < 0.001 | 9,689 | 870 | 10 |
| KDM7 | Respondent-destination rank: +10 percentiles | Unit weights | -0.032 | -0.039, -0.025 | < 0.001 | 9,797 | 9,797 | 11 |

Continued on next page

Table S11 continued

| KDM | Exposure contrast | Weighting scheme | Est. | 95% CI | P | n | ESS | Countries |
|---|---|---|---|---|---|---|---|---|
| KDM5 | Respondent education: +1 weighted SD | Official DBS | -0.076 | -0.124, -0.029 | 0.002 | 14,015 | 1,867 | 12 |
| KDM5 | Respondent education: +1 weighted SD | Trimmed official DBS | -0.077 | -0.125, -0.029 | 0.002 | 14,015 | 1,878 | 12 |
| KDM5 | Respondent education: +1 weighted SD | Trimmed SIPW | -0.088 | -0.140, -0.036 | < 0.001 | 14,015 | 1,555 | 12 |
| KDM5 | Respondent education: +1 weighted SD | Unit weights | -0.103 | -0.121, -0.085 | < 0.001 | 14,015 | 14,015 | 12 |
| KDM5 | Highest-parent education: +1 weighted SD | Official DBS | -0.072 | -0.114, -0.030 | < 0.001 | 12,974 | 1,773 | 11 |
| KDM5 | Highest-parent education: +1 weighted SD | Trimmed official DBS | -0.071 | -0.113, -0.030 | < 0.001 | 12,974 | 1,785 | 11 |
| KDM5 | Highest-parent education: +1 weighted SD | Trimmed SIPW | -0.086 | -0.131, -0.040 | < 0.001 | 12,974 | 1,435 | 11 |
| KDM5 | Highest-parent education: +1 weighted SD | Unit weights | -0.064 | -0.081, -0.046 | < 0.001 | 12,974 | 12,974 | 11 |
| KDM5 | Wealth rank: +10 percentiles | Official DBS | -0.072 | -0.089, -0.055 | < 0.001 | 13,961 | 1,852 | 12 |
| KDM5 | Wealth rank: +10 percentiles | Trimmed official DBS | -0.073 | -0.090, -0.056 | < 0.001 | 13,961 | 1,863 | 12 |
| KDM5 | Wealth rank: +10 percentiles | Trimmed SIPW | -0.072 | -0.091, -0.054 | < 0.001 | 13,961 | 1,542 | 12 |
| KDM5 | Wealth rank: +10 percentiles | Unit weights | -0.063 | -0.070, -0.056 | < 0.001 | 13,961 | 13,961 | 12 |
| KDM5 | Income rank: +10 percentiles | Official DBS | -0.065 | -0.081, -0.048 | < 0.001 | 12,383 | 1,745 | 12 |
| KDM5 | Income rank: +10 percentiles | Trimmed official DBS | -0.066 | -0.082, -0.049 | < 0.001 | 12,383 | 1,758 | 12 |
| KDM5 | Income rank: +10 percentiles | Trimmed SIPW | -0.060 | -0.079, -0.041 | < 0.001 | 12,383 | 1,404 | 12 |
| KDM5 | Income rank: +10 percentiles | Unit weights | -0.054 | -0.060, -0.047 | < 0.001 | 12,383 | 12,383 | 12 |
| KDM5 | Residualized educational-rank mobility: +1 weighted SD | Official DBS | -0.083 | -0.127, -0.040 | < 0.001 | 12,974 | 1,773 | 11 |
| KDM5 | Residualized educational-rank mobility: +1 weighted SD | Trimmed official DBS | -0.085 | -0.128, -0.041 | < 0.001 | 12,974 | 1,785 | 11 |
| KDM5 | Residualized educational-rank mobility: +1 weighted SD | Trimmed SIPW | -0.088 | -0.134, -0.042 | < 0.001 | 12,974 | 1,435 | 11 |
| KDM5 | Residualized educational-rank mobility: +1 weighted SD | Unit weights | -0.097 | -0.115, -0.080 | < 0.001 | 12,974 | 12,974 | 11 |
| KDM5 | Parental-origin rank: +10 percentiles | Official DBS | -0.013 | -0.029, 0.004 | 0.135 | 12,974 | 1,773 | 11 |
| KDM5 | Parental-origin rank: +10 percentiles | Trimmed official DBS | -0.013 | -0.029, 0.004 | 0.134 | 12,974 | 1,785 | 11 |

Continued on next page

Table S11 continued

| KDM | Exposure contrast | Weighting scheme | Est. | 95% CI | P | n | ESS | Countries |
|---|---|---|---|---|---|---|---|---|
| KDM5 | Parental-origin rank: +10 percentiles | Trimmed SIPW | -0.017 | -0.035, -0.000 | 0.048 | 12,974 | 1,435 | 11 |
| KDM5 | Parental-origin rank: +10 percentiles | Unit weights | -0.009 | -0.016, -0.002 | 0.012 | 12,974 | 12,974 | 11 |
| KDM5 | Respondent-destination rank: +10 percentiles | Official DBS | -0.031 | -0.047, -0.014 | $< 0.001$ | 12,974 | 1,773 | 11 |
| KDM5 | Respondent-destination rank: +10 percentiles | Trimmed official DBS | -0.031 | -0.048, -0.015 | $< 0.001$ | 12,974 | 1,785 | 11 |
| KDM5 | Respondent-destination rank: +10 percentiles | Trimmed SIPW | -0.032 | -0.050, -0.015 | $< 0.001$ | 12,974 | 1,435 | 11 |
| KDM5 | Respondent-destination rank: +10 percentiles | Unit weights | -0.036 | -0.043, -0.030 | $< 0.001$ | 12,974 | 12,974 | 11 |

*Note.* Estimates are differences in KDM age acceleration in SD units. ESS denotes weighted effective sample size. The table displays the weighting schemes used in the point-estimate sensitivity comparison. Raw KDM5 SIPW showed greater weight instability (maximum multiplier, 36.40); the KDM5 point-estimate sensitivity comparison therefore includes trimmed SIPW rather than raw SIPW.

**Table S12:** Selection-bootstrap availability

| KDM | Estimand or test | Weighting scheme | Available | Expected | Fraction |
|---|---|---|---|---|---|
| KDM9 | Pooled coefficients | SIPW | 7 | 7 | 1.00 |
| KDM9 | Pooled coefficients | Trimmed SIPW | 7 | 7 | 1.00 |
| KDM9 | Pooled joint tests | SIPW | 6 | 6 | 1.00 |
| KDM9 | Pooled joint tests | Trimmed SIPW | 6 | 6 | 1.00 |
| KDM9 | Country-interaction coefficients | SIPW | 66 | 73 | 0.90 |
| KDM9 | Country-interaction coefficients | Trimmed SIPW | 66 | 73 | 0.90 |
| KDM9 | Country-interaction joint tests | SIPW | 0 | 6 | 0.00 |
| KDM9 | Country-interaction joint tests | Trimmed SIPW | 0 | 6 | 0.00 |
| KDM9 | Country-specific slopes | SIPW | 73 | 84 | 0.87 |
| KDM9 | Country-specific slopes | Trimmed SIPW | 73 | 84 | 0.87 |
| KDM7 | Pooled coefficients | SIPW | 7 | 7 | 1.00 |
| KDM7 | Pooled coefficients | Trimmed SIPW | 7 | 7 | 1.00 |
| KDM7 | Pooled joint tests | SIPW | 6 | 6 | 1.00 |
| KDM7 | Pooled joint tests | Trimmed SIPW | 6 | 6 | 1.00 |
| KDM7 | Country-interaction coefficients | SIPW | 66 | 66 | 1.00 |
| KDM7 | Country-interaction coefficients | Trimmed SIPW | 66 | 66 | 1.00 |
| KDM7 | Country-interaction joint tests | SIPW | 6 | 6 | 1.00 |
| KDM7 | Country-interaction joint tests | Trimmed SIPW | 6 | 6 | 1.00 |
| KDM7 | Country-specific slopes | SIPW | 73 | 84 | 0.87 |
| KDM7 | Country-specific slopes | Trimmed SIPW | 73 | 84 | 0.87 |
| KDM5 | Pooled coefficients | Trimmed SIPW | 7 | 7 | 1.00 |
| KDM5 | Pooled joint tests | Trimmed SIPW | 6 | 6 | 1.00 |
| KDM5 | Country-interaction coefficients | Trimmed SIPW | 66 | 73 | 0.90 |
| KDM5 | Country-interaction joint tests | Trimmed SIPW | 0 | 6 | 0.00 |
| KDM5 | Country-specific slopes | Trimmed SIPW | 73 | 84 | 0.87 |

*Note.* A bootstrap uncertainty estimate was considered available when at least 950 of 1,000 replicates completed successfully. Fraction is the number of estimates or tests meeting this criterion divided by the number expected.

**Table S13:** Sources and extraction definitions for the six country-level moderators

| Moderator | Source and locator | Reference period and unit | Selection and transformation |
|---|---|---|---|
| Public social expenditure | OECD SOCX SDMX extract [8]; dataflow DSD_SOCX_AGG@DF_SOCX_AGG, version 1.0 | 2015; % of GDP | Annual SOCX; unit PT_B1GQ; public expenditure ES10; total spending and programme types (SPENDING_TYPE=_T, PROGRAMME_TYPE=_T). |
| Disposable-income Gini | OECD IDD SDMX extract [9]; dataflow DSD_WISE_IDD@DF_IDD, version 1.0 | 2015; coefficient on the 0–1 scale | Annual INC_DISP_GINI; all ages (AGE=_T); METHODOLOGY=METH2012; UNIT_MEASURE=0_TO_1. Definition selection is specified below. |
| Low-earner net pension replacement | OECD *Pensions at a Glance 2015* [10]; Table 6.7, p. 145 (revised March 2016) | 2014 policy rules; net replacement rate, % | Earnings at 0.5 times average earnings. Male and female rates were averaged with equal weights; where the table reported no sex difference, the common rate was retained. |
| Out-of-pocket health expenditure | WHO GHED via WDI [11, 12]; SH.XPD.OOPC.CH.ZS | 2015; % of current health expenditure | Total out-of-pocket expenditure as a share of current health expenditure. |
| Log GDP per capita, PPP | World Bank WDI [12]; NY.GDP.PCAP.PP.KD | 2015; constant 2021 international dollars | The natural logarithm of GDP per capita at purchasing-power parity was taken before standardization. |
| Unemployment | ILO modeled estimates via WDI [12]; SL.UEM.TOTL.ZS | 2015; % of total labor force | Total unemployment, both sexes, using the modeled ILO estimate. |

*Note.* All source files were retrieved on 23 August 2026. WDI and OECD IDD queries covered 2014–2016; the analysis selected the exact 2015 observations for all 12 countries. The pension measure instead used the 2014 policy rules in the cited report. For eligible OECD Gini records, definition flags were selected in the order D_CUR, then D_INC, then D_PREV. Belgium and Israel used D_INC, France used D_PREV, and the other nine countries used D_CUR. Each moderator was standardized by subtracting its equal-country mean and dividing by its sample standard deviation across the same 12 countries; GDP was logged first. The hyperlinks identify the OECD extraction queries, pension report, and WDI indicator pages.
